# Linguistic Effects of Ambient AI on Clinical Documentation: A Matched Pre-Post Study

**DOI:** 10.64898/2026.02.16.26346370

**Authors:** Yiming Li, Huiyuan Zhou, Suzanne Blackley, Joseph M. Plasek, Zhuoyang Lyu, Wei Zhang, Jacqueline You, Amanda Centi, Rebecca Mishuris, Jie Yang, Li Zhou

## Abstract

Ambient intelligence–based systems are increasingly used for clinical documentation. To quantify linguistic differences associated with ambient documentation, we conducted a matched pre–post analysis of 6,026 outpatient clinical notes from Mass General Brigham following implementation of two ambient AI documentation systems (Nuance Dragon Ambient eXperience [DAX] and Abridge). Within-clinician comparisons focused on the History of Present Illness (HPI) and Assessment and Plan (A&P) sections and evaluated syntactic complexity, lexical ambiguity, linguistic variability, discourse coherence, and readability. Manual review of 50 paired notes was performed to validate findings from automated linguistic analyses.

Our analyses indicate that the linguistic effects of ambient documentation are both vendor-dependent and section-specific. Across both vendors, ambient notes in HPI were longer and exhibited greater syntactic complexity (longer sentences and clauses, increased dependency distance), lower lexical ambiguity, lower language-model perplexity, and higher local and global discourse coherence. These findings indicate that ambient systems systematically restructure conversational input into more syntactically elaborated and linguistically predictable narratives, reflecting increased standardization relative to both general-domain and biomedical language models. In contrast, changes in A&P were smaller and more heterogeneous, consistent with its more structured/templated nature. Readability analyses further showed increased length and lexical complexity in ambient HPI, whereas A&P readability differences were minimal.

Overall, our findings demonstrate that ambient documentation changes how clinical information is linguistically expressed and organized, with effects varying by note section, vendor, and provider role/specialty. Evaluation should therefore extend beyond efficiency to consider effects on communication, cognitive load, clinical inference, and downstream analytics.

## 1. Introduction

Ambient intelligence systems that automatically generate clinical notes from provider–patient conversations using speech recognition and large language models (LLMs) represent a milestone revolution in medical documentation [1]. These systems aim to reduce the clerical burden, improve workflow efficiency, and enable clinicians to devote more time to patient care [2]. In a quality improvement study, Stults et al. reported that implementation of an ambient intelligence (AI) documentation platform reduced mean time spent on charting from 6.2 to 5.3 minutes per visit, resulting in lower cognitive load [3]. Similarity, in a survey of 1,430 clinicians, You et al. found that adoption of ambient documentation technology was associated with reductions in reported burnout (from 50.6% to 29.4%) and improved documentation-related well-being [4], [5], [6].

In addition to workflow improvements, ambient systems may also alter the linguistic properties of clinical documentation, with potential effects on clarity and interpretability. Unlike traditional approaches such as dictation and medical scribes, ambient systems rely on LLMs that are typically pre-trained on broad, predominantly general-domain corpora and subsequently adapted through domain-specific fine-tunings [7], [8]. Because clinical documentation represents a specialized sublanguage, pre-training data may provide limited exposure to its semantic, terminological, and discourse conventions. Although fine-tuning and instruction-based prompting aim to improve clinical alignment, they may not fully capture the heterogeneity of real-world scenarios, specialty-specific language, evolving practice patterns, or institution-level documentation norms. Consequently, generated notes may read fluently but often differ from clinician-authored notes in terms of structural organization, lexical specificity, or discourse coherence [9], [10]. Even subtle shifts in syntactic structure, terminology, or information sequencing could influence clinician interpretation, cognitive load, and trust in AI-generated documentation [11] [12] [13], [14].

High-quality clinical notes are essential for accurate communication, continuity of care, and downstream applications such as clinical decision support, billing, and secondary data analysis [12]. As notes are increasingly shared with patients through electronic portals, clarity and readability may also affect patient understanding and engagement. Linguistic features, including sentence complexity, coherence, and ambiguity, directly affect how effectively clinical meaning is conveyed [13], [14]. Whereas clinician-authored notes demonstrate individual narrative and reasoning style, ambient-generated notes are shaped by learned generative patterns and constrained by systemic factors such as model architecture, training data characteristics, and preconfigured documentation templates, potentially affecting the balance of information across sections, lexical specificity, and discourse coherence [15], [16]. Qualitative findings by Tiem et al. supported these concerns, reporting perceived limitations in ambient-assisted notes, including overlong or underspecified sections, unfamiliar formatting, and diminished clinical “voice” [17].

Despite growing adoption, most evaluations of ambient documentation systems have focused on efficiency, time savings, and user satisfaction rather than objective linguistic quality [3], [18], [19]. Early studies frequently relied on subjective user feedback without quantitative language analysis[20], [21]. Separately, prior research on traditional clinical documentation has demonstrated substantial variability in readability and structure across sections such as the History of Present Illness (HPI) and Assessment and Plan (A&P) [22], [23], [24], [25]. Established metrics such as Flesch-Kincaid Grade Level (FKGL), syntactic complexity, and lexical diversity have been used to assess documentation clarity and its implications for comprehension and clinical communication [26], [27]. More recent work has applied natural language processing (NLP) and LLMs to assess coherence, ambiguity, and contextual clarity in electronic health record (EHR) text [28], [29]. However, few studies have systematically compared ambient-generated and clinician-authored notes across these linguistic dimensions, leaving a critical gap in understanding how ambient intelligence influences the clarity and interpretability of clinical documentation.

To address this gap, we conducted a comparative analysis of ambient-assisted and clinician-authored notes from the HPI and A&P sections. Notes were evaluated across five dimensions: syntactic complexity, lexical ambiguity, linguistic variability, discourse coherence, and readability. We additionally compared outputs from two ambient documentation vendors to examine whether system design influences linguistic characteristics. By quantitatively assessing documentation across multiple linguistic dimensions, this study seeks to clarify where systematic differences emerge and to guide future research on the evaluation and responsible integration of ambient documentation systems in clinical practice.

## 2. Methods

This study employed a multidimensional linguistic analysis to compare ambient-assisted and clinician-authored notes across several dimensions of textual quality (Figure 1). Specifically, notes were evaluated across four domains: 1) **Syntactic Complexity**, quantified using metrics such as sentence length, clause length, and dependency distance; 2) **Lexical Ambiguity**, examined at WordNet-based sense enumeration; 3) **Linguistic Variability**, measured with language model perplexity 4) **Discourse Coherence**, evaluated at both local and global levels using embedding-based semantic similarity measures; 5) **Readability**, assessed using the validated MedReadMe approach. Collectively, these dimensions characterize structural and semantic properties of clinical text that may influence clarity, interpretability, cognitive processing, and reliability for both direct patient care and secondary data use. We further compared these linguistic measures across vendor systems, note sections, and provider characteristics (provider type, specialty and sex).

**Figure 1.**
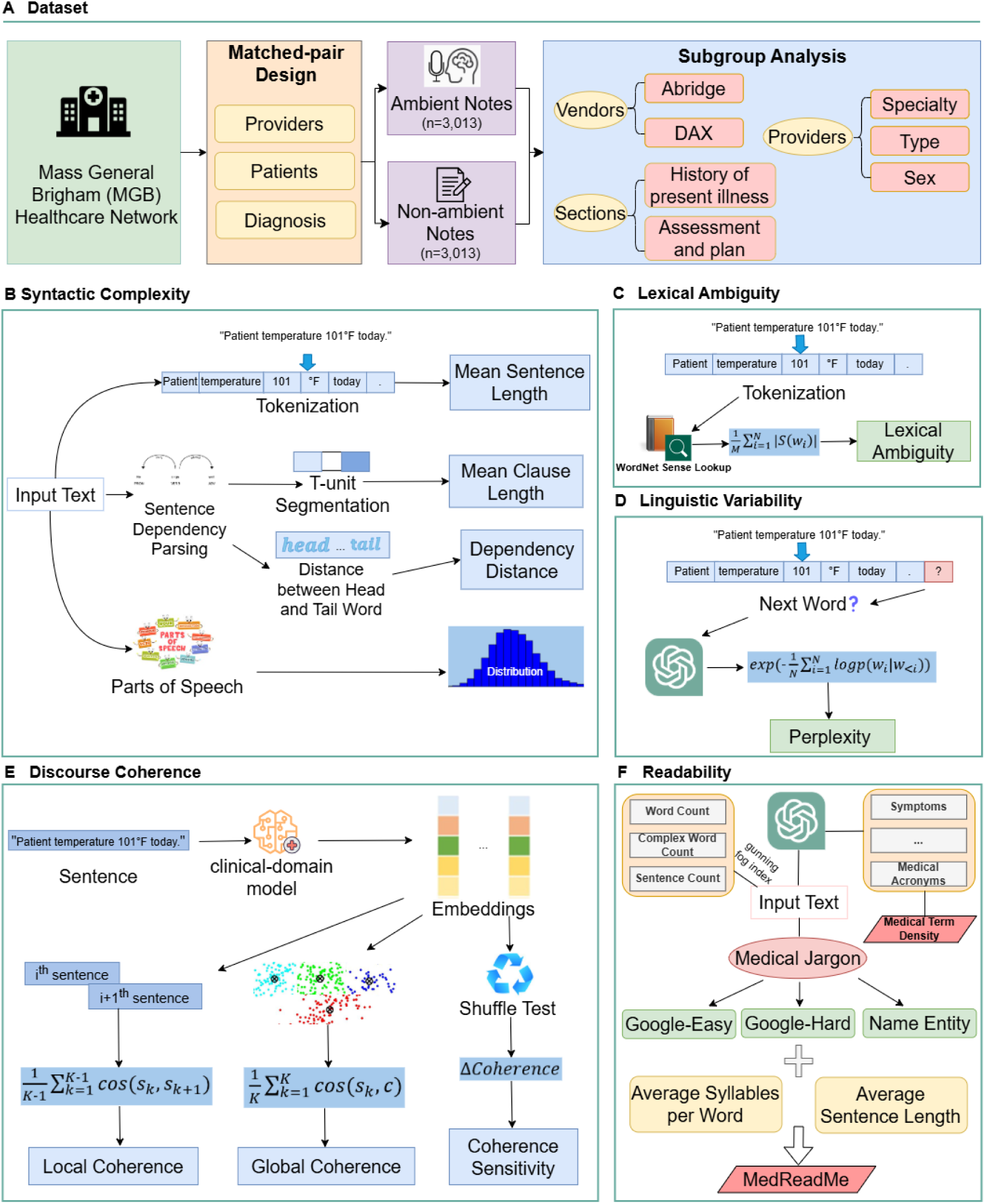
Overview of the dataset and analytical framework for comparing ambient and non-ambient clinical notes. (A) **Dataset construction.** Clinical notes were extracted from the Mass General Brigham (MGB) electronic health record database. Ambient and non-ambient cohorts were constructed by identifying notes from the same provider–patient pairs matched on primary encounter diagnosis. Linguistic measures were further compared across vendor systems, note sections, and provider characteristics. (B-E) illustrate four major linguistics dimensions. (B) **Syntactic complexity.** Text was processed using tokenization, sentence and T-unit segmentation, dependency parsing, and part-of-speech tagging to compute mean sentence length, mean clause length, dependency distance, and part-of-speech distributions. (C) **Lexical Ambiguity.** Lexical ambiguity was measured via WordNet sense lookup, following tokenization and ambient/non-ambient classification. (D) **Linguistic Variability**. Semantic ambiguity was evaluated via language-model–based perplexity. (E) **Discourse Coherence**. Local and global coherence were estimated using sentence embeddings derived from clinical-domain models, with coherence sensitivity assessed via a shuffle test. (F) **Readability**. Readability was quantified using MedReadMe Framework, which provides fine-grained, sentence-level readability assessment for medical text. The analysis incorporates medical jargon features classified as “Google-Easy,” “Google-Hard,” and “Named Entity” (e.g., brand or organization names), in addition to average sentence length and average syllables per word to compare ambient versus non-ambient notes.

### 2.1. Dataset

This study was conducted within Mass General Brigham (MGB), a large integrated healthcare network based in Boston, Massachusetts. At the time of analysis, MGB was piloting two ambient clinical documentation systems (Nuance Dragon Ambient eXperience [DAX] and Abridge) across multiple outpatient sites.

We extracted all ambulatory office visit notes across medical and surgical specialties generated using ambient documentation between January 1 and March 31, 2025, to form the ambient dataset. For comparison, we retrieved office visit notes authored by the same provider for the same patient and primary encounter diagnosis between January 1 and March 31, 2023, prior to implementation of ambient documentation at MGB (non-ambient dataset). In total, 3,013 unique provider–patient pairs were identified. When multiple notes were available for a given pair within a study period, one note was randomly selected from the ambient period and one from the pre-implementation period, yielding a final sample of 6,026 clinical notes for analysis. The demographic characteristics of the patients and providers are summarized in Supplementary Table 1. All procedures were reviewed and approved by the Mass General Brigham Institutional Review Board (IRB).

### 2.2 Syntactic Complexity

#### 2.2.1 Mean Sentence and Clause Length

Sentence and clause length were analyzed as indicators of intra-sentential syntactic complexity and expressive density in clinical documentation. These measures characterize how information is structured within sentences and may influence processing effort and interpretability. Longer syntactic units can encode more information within a single construction but may also increase cognitive load and reduce clarity.

Mean sentence length was calculated as the average number of word tokens per sentence following standard sentence segmentation. This metric provides a general and widely used index of structural complexity in readability and stylistic analyses. Very short sentences may indicate fragmentary or telegraphic writing, whereas overly long sentences may reduce clarity due to heavy coordination or clause embedding.

To capture syntactic complexity at a finer level, we further analyzed clause length using T-units, defined as the smallest grammatically complete units, consisting of one independent (main) clause and all of its dependent subordinate clauses. For example, “The patient improved” is a one T-unit, and “The patient improved after receiving antibiotics” is still a one T-unit because the subordinate clause depends on the main clause. However, “The patient improved after receiving antibiotics, and the fever resolved within 24 hours” contains two T-units, as each clause could stand alone. T-unit segmentation was implemented using syntactic parsing outputs derived from the spaCy dependency parser to ensure consistent identification of clause boundaries. Mean T-unit length (MTUL) was computed for each note as:

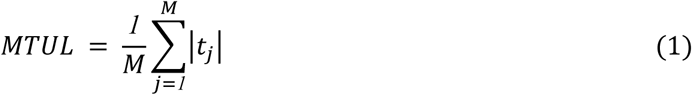

where 𝑀 is the total number of T-units in the note, and |𝑡_𝑗_| denotes the number of words in the 𝑗^𝑡*ℎ*^ T-unit. Both mean sentence length and mean T-unit length were calculated on a per-note basis to enable comparison across documents of varying length. These two complementary measures jointly quantify syntactic elaboration, distinguishing multi-clausal, information-dense constructions from simpler sentence structures.

#### 2.2.2 Dependency Distance

Dependency distance was employed to quantify syntactic complexity and structural cohesion of clinical text [30]. This metric measures the average linear distance between syntactically linked words, typically a head and its dependent within a sentence. Shorter distances generally reflect locally organized structures, whereas longer distances indicate greater hierarchical embedding and structural dispersion, potentially increasing processing demands for both human readers and NLP systems.

For example, in the sentence *“The patient denies chest pain,”* the dependency between the subject *patient* and the verb *denies* spans only a short linear distance, reflecting a compact syntactic structure. In contrast, in *“The patient who presented to the emergency department last night denies chest pain,”* the subject *patient* and the verb *denies* are separated by an intervening relative clause. This increases their linear distance and illustrates how syntactic embedding contributes to greater dependency distance.

Each sentence was parsed using the spaCy dependency parser (version 3.8.7), which identifies grammatical relationships between tokens following Universal Dependencies conventions. For each dependency relation between a head word *ℎ*_𝑖_ and its dependent 𝑑_𝑖_, the absolute difference between their token positions was computed. Mean dependency distance (MDD) for a sentence was defined as:

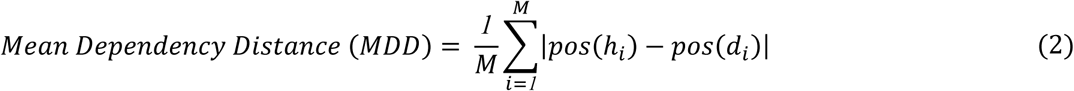

where 𝑝𝑜𝑠(⋅) denotes the token position within the sentence, and 𝑀 represents the total number of dependency relations. Because MDD is computed as the mean distance across all dependencies in a sentence, it inherently adjusts for variation in sentence length. Sentence-level MDD values were averaged across sentences within each note to obtain a document-level measure.

Dependency distance distributions were then compared between ambient and non-ambient cohorts to evaluate whether automated documentation systems influence syntactic cohesion and overall structural complexity in clinical narratives.

#### 2.2.3 Part-of-Speech (POS) Distribution

Part-of-speech analysis was conducted to examine the lexical and grammatical composition of clinical notes and to identify stylistic differences between ambient and non-ambient documentation. The distribution of POS categories (e.g., nouns, verbs, adjectives, and adverbs) reflects how information is structured and functionally expressed within a text. Variations in POS profiles may indicate differences in descriptive density, action-oriented phrasing, or narrative style, which can affect clarity, informativeness, and readability.

POS tagging was conducted using the spaCy v3.8.7 NLP library, which employs a transition-based dependency parser and pre-trained statistical model optimized for biomedical and clinical text. Each token in a note was assigned a Universal POS tag, allowing consistent comparison across corpora. Tokens such as punctuation and special characters were excluded from analysis to avoid artificial inflation of counts.

For each note, the relative frequency of major POS categories was calculated as:

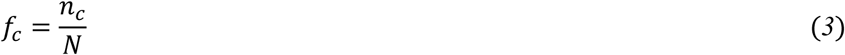

where 𝑛_𝑐_ denotes the number of tokens belonging to category 𝑐, and 𝑁 is the total number of tokens in the note.

In addition to POS category frequencies, POS entropy was calculated to quantify the overall diversity and balance of part-of-speech usage within each note. POS entropy was defined using Shannon’s entropy formulation:

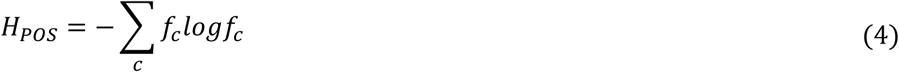

where 𝑓_𝑐_ is the relative frequency of POS category 𝑐. Higher POS entropy indicates a more diverse and evenly distributed use of grammatical categories, whereas lower entropy reflects a more concentrated or uniform grammatical structure.

### 2.3 Lexical Ambiguity (Polysemy-based Measure)

Lexical ambiguity was assessed at the word level to quantify variability in potential word meaning within clinical documentation. Ambiguous terms may reduce interpretability and increase the risk of miscommunication. We operationalized lexical ambiguity as polysemy, i.e., the number of distinct senses associated with a word in a standardized lexical-semantic resource [31]. Document-level lexical ambiguity was measured as the mean number of WordNet senses per token:

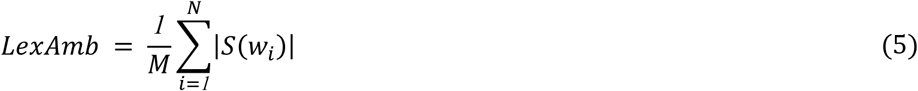

where 𝑆(𝑤_𝑖_) denotes the set of WordNet senses associated with word 𝑤_𝑖_, |𝑆(𝑤_𝑖_)| is the number of senses for that word, and 𝑁 is the total number of tokens in the document.

WordNet provides structured lists of English word meanings, separating the different senses associated with a single word. For example, *“discharge”* has distinct senses referring to releasing a patient, emitting fluid, or settling a financial obligation, each represented separately in WordNet. Although WordNet is not specific to clinical terminology, it offers a consistent and reproducible proxy for estimating lexical polysemy across documents. Higher LexAmb values indicate greater average sense inventory size and thus greater potential lexical ambiguity. Lower values reflect few available senses per word and may correspond to more semantically constrained vocabulary. Note that this metric captures ambiguity at the level of sense inventory size rather than contextual ambiguity within discourse.

### 2.4. Linguistic Variability (Perplexity-based Approximation)

Linguistic variability refers to the degree to which a text deviates from the distributional patterns learned by a language model. It is operationalized as model-relative unpredictability, quantified using perplexity. Perplexity quantifies the predictability of a token sequence under a probabilistic language model [32], reflecting how well preceding context predicts subsequent tokens. Text that aligns closely with a model’s learned distribution yields lower perplexity, whereas text that deviates from typical lexical or structural patterns relative to that model produces higher perplexity [33].

Perplexity was calculated using both a general-domain GPT-2 model and a domain-specific BioGPT model to account for potential differences in medical language modeling. For a text sequence 𝑇 consisting of 𝑁 tokens, perplexity is defined as:

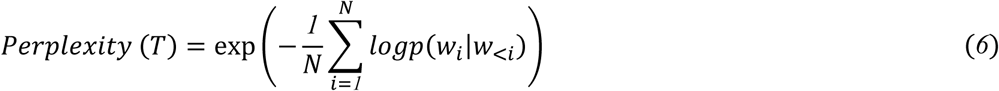

where 𝑤_𝑖_ represents the 𝑖^𝑡^*^ℎ^*token, and 𝑝(𝑤_𝑖_|𝑤_<𝑖_) denotes the conditional probability of that token given preceding context.

Perplexity was calculated using each model’s native tokenizer and vocabulary. Higher perplexity values indicate lower predictability relative to the model’s learned distribution. Comparing GPT-2 and BioGPT perplexity enables differentiation between general linguistic atypicality and deviations specific to biomedical language usage.

### 2.5 Discourse Coherence

Discourse coherence analysis was conducted to evaluate the logical and semantic continuity of clinical documentation [34]. Coherence indicates how effectively sentences connect to convey a unified narrative. This analysis can help determine if AI-generated ambient notes maintain the structural integrity and thematic focus or if they exhibit "hallucinated" transitions and stylistic fragmentation.

#### 2.5.1 Local Coherence

Local coherence measures the semantic relatedness between adjacent sentences, capturing the smoothness of transitions and the continuity of micro-level thought. Each sentence was embedded using Bio_ClinicalBERT [35], to capture domain-specific semantic nuances. The local coherence score was defined as the mean cosine similarity between consecutive sentence embeddings:

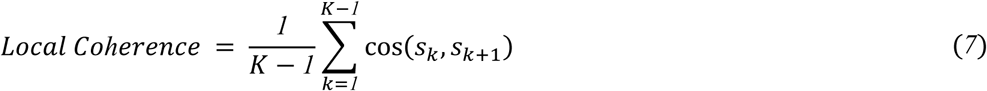

where 𝑠_𝑘_ and 𝑠_𝑘+_*_1_* denote the embeddings of the 𝑘-th and (𝑘 + *1*)-th sentences, and 𝐾 is the total number of sentences in the note. Higher values indicate smoother conceptual transitions, while lower values suggest abrupt topic shifts or logical "jumps" that may impede clinical comprehension.

#### 2.5.2 Global Coherence

Global coherence assesses macro-level thematic consistency by quantifying how closely each sentence aligns with the note’s overarching context. To represent this context, a document centroid vector (*c*) was calculated as the mean of all sentence embeddings. Global coherence is defined as the average cosine similarity between each sentence and this centroid:

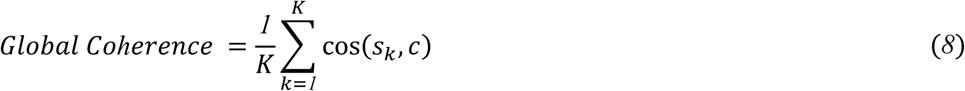

where 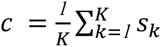 represents the document’s semantic centroid. High global coherence reflects unified thematic focus, whereas lower scores suggest narrative fragmentation or tangential content.

#### 2.5.3 Coherence Sensitivity (Shuffle Test)

To validate the sensitivity and robustness of coherence metrics, we employed a shuffle-based permutation test. Because coherence is inherently dependent on linear order, randomly permuting sentence sequence should significantly degrade the scores. The resulting difference between original and shuffled scores (𝛥𝐶𝑜*ℎ*𝑒𝑟𝑒𝑛𝑐𝑒) was computed as:

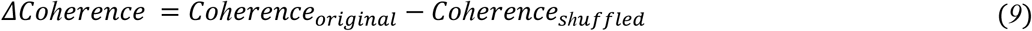

A larger 𝛥𝐶𝑜*ℎ*𝑒𝑟𝑒𝑛𝑐𝑒 denotes stronger discrimination between ordered and disordered text, confirming that the embeddings effectively encode the underlying discourse structure.

By combining these different measurements, we can accurately judge if AI-generated notes stay logically organized and easy for readers to follow, or if they become fragmented and confusing.

### 2.6 Readability

Readability was assessed to estimate the cognitive effort required to interpret clinical notes. As a preliminary step, we computed basic text statistics using the Gunning Fog Index framework, including total word count, sentence count, and complex word count (defined as words containing three or more syllables). These components were used to characterize lexical-level textual complexity [36].

To explicitly quantify the use of domain-specific terminology, we additionally measured Medical Term Density. Medical terms were extracted using GPT-4o and categorized into clinically meaningful classes, including Signs and Symptoms, Diseases and Diagnoses, Medications, Procedures, Laboratory Tests, Radiology and Imaging Studies, Vital Signs and Clinical Measurements, Lifestyle and Social History Factors, Family History, Social Determinants of Health, and Medical Acronyms. Medical term density was defined as the proportion of medical terms within a clinical text, reflecting the concentration of domain-specific vocabulary. Formally, for a given text containing 𝑁 total tokens, medical term density was calculated as Formula (10):

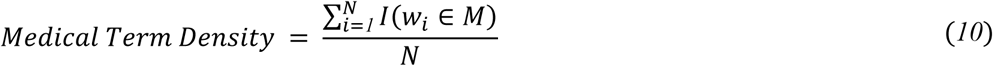

where 𝑤_𝑖_ denotes the 𝑖-th token in the text, 𝑀 represents the medical terminology, and 𝐼(.) is an indicator function that equals 1 if the token is identified as a medical term and 0 otherwise.

Because general-purpose readability formulas (e.g., Flesch–Kincaid) do not adequately capture medical terminology and domain-specific complexity, we employed MedReadMe [31], [37], a clinically tailored readability framework developed from a manually annotated corpus of medical sentences. MedReadMe incorporates domain-relevant features, including medical jargon and span-level complexity annotations, and has demonstrated stronger alignment with human judgments of difficulty than traditional readability metrics in clinical text.

We applied the best-performing MedReadMe model from the published repository to generate readability scores for each note [32]. MedReadMe extends traditional readability metrics by explicitly incorporating medical jargon. A weighted jargon term was added to standard scores (e.g., FKGL), with weights tuned via grid search on a development set using gold-standard annotations. Jargon spans were identified using the proposed best-performing complex span identification model, and incorporating this term substantially improved correlations with human judgments of readability. Higher scores indicate greater linguistic complexity and expected processing effort, whereas lower scores reflect more accessible language. Comparing readability distributions between ambient and non-ambient notes enables assessment of whether automated documentation systems systematically alter interpretability in clinical narratives.

### 2.7 Manual Evaluation

We conducted a manual evaluation to assess the coherence and readability of clinical notes. Two independent physician evaluators rated the notes using a 5-point Likert scale (1 = lowest, 5 = highest). A total of 50 paired clinical notes were randomly selected, consisting of notes created before and after ambient use for the same provider-patient pair. Among the ambient notes, 25 pairs were generated using ambient documentation technology from Vendor I, and the remaining 25 were generated using Vendor II.

Both evaluators independently assessed the HPI and A&P sections of all clinical notes. The evaluators were blinded to the documentation method and vendor. Scores were averaged and aggregated for downstream statistical analysis.

### 2.8 Statistical Analysis

We evaluated differences in linguistic characteristics between ambient and non-ambient clinical notes across multiple outcome domains, including syntactic complexity, lexical ambiguity, linguistic predictability, coherence, and readability, as described in the Methods. Analyses were conducted separately for the HPI and A&P sections and were additionally stratified by vendor system.

The analytic dataset consisted of patients nested within providers, with each patient contributing exactly two observations: one non-ambient note and one ambient note. For each outcome, we summarized results using descriptive statistics (mean [SD]) for both notes. To quantify the overall ambient-associated changes in linguistic features, we calculated the within-patient difference by subtracting the non-ambient value from the ambient value for each outcome. Because each patient served as their own control and did not switch providers over time, the paired design inherently accounted for all time-invariant patient- and provider-level characteristics. No covariate adjustment was needed for estimation of the overall mean change. Intercept-only linear regression models were then fitted, where the intercept represents the mean ambient vs. non-ambient difference for each linguistic measure.

Provider characteristics were harmonized prior to modeling to ensure meaningful subgroup comparisons. Specifically, for provider specialty, we grouped “internal medicine/primary care” and “pediatrics” into primary care; “general surgery,” “orthopedics,” “otolaryngology,” “thoracic surgery,” “urology,” and “OB/GYN” into “surgical subspecialty”; and all remaining specialties into medical subspecialty. For provider type, we grouped “MD,” “DO,” “MBBS,” and “MBBCH” as physician, and all other credentials as non-physician [38]. To explore whether ambient-associated changes differed according to provider characteristics, multivariable linear regression models were fitted by regressing difference scores on provider characteristics including sex, specialty, and type. Adjusted marginal mean changes and corresponding confidence intervals for each level of the patient characteristics were estimated using the ‘*emmeans*’ package.

For manual evaluation, inter-rater agreement between the two evaluators was assessed using a two-way intraclass correlation coefficient (ICC). To compare coherence and readability of ambient vs. non-ambient clinical notes by physician evaluators, we first assessed the normality of paired differences using the Shapiro–Wilk test. Paired *t* tests were applied for metrics with normally distributed differences; otherwise, the Wilcoxon signed-rank test was used. To evaluate potential systematic differences between manual evaluation scores and algorithm-generated scores, all scores were standardized using Z-score transformation, and the same statistical testing procedure was employed.

To account for the potential correlation among patients treated by the same provider, we applied Huber–White heteroskedasticity-robust standard errors clustered at the provider level for all regression analyses. All statistical analyses were performed using R version 4.4.1 (R Foundation for Statistical Computing) and two-sided *P* value < .05 was considered as statistically significant.

## 3. Results

Across both vendors, mean differences between non-ambient and ambient notes showed a section-specific pattern. For Vendor I, ambient documentation in the HPI was associated with higher mean sentence length (17.96 vs 16.61 words), greater mean clause length (14.12 vs 12.69 words per T-unit), and higher dependency distance (3.00 vs 2.84 words), suggesting increased syntactic complexity in this section. In contrast, the A&P section showed the opposite direction: mean sentence length (20.52 vs 25.81), mean clause length (15.34 vs 18.35), and dependency distance (3.09 vs 3.34) were all lower in ambient notes compared with non-ambient notes, indicating relatively simpler syntactic structure.

For Vendor II, mean differences were smaller in magnitude. In the HPI, ambient notes had modestly higher mean sentence length (18.46 vs 17.86), clause length (14.14 vs 13.39), and dependency distance (2.94 vs 2.88), again suggesting slightly greater complexity. However, in the A&P section, mean sentence length was nearly unchanged (22.60 vs 22.77), while clause length (16.56 vs 16.97) and dependency distance (3.13 vs 3.19) were slightly lower in ambient notes. Overall, the mean findings suggest that ambient documentation tends to increase syntactic complexity in the HPI but has minimal or slightly simplifying effects in the A&P, with more pronounced differences observed for Vendor I.

**Table 1.**
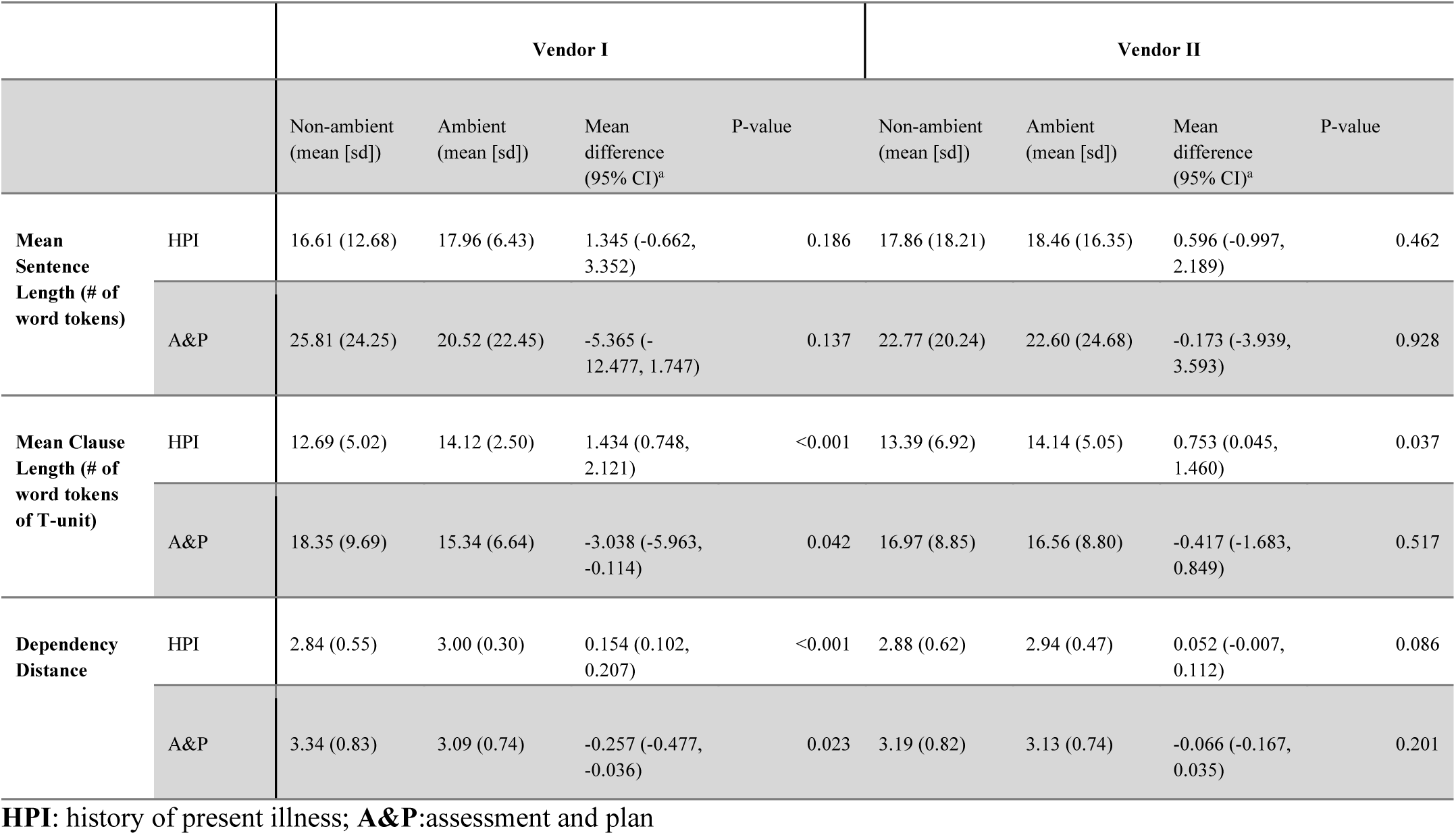
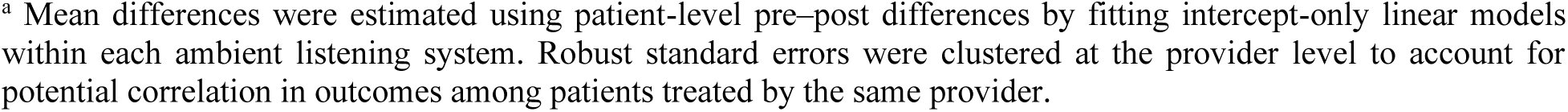
Differences in syntactic complexity between ambient and non-ambient clinical notes by note section and vendor.

|  |  | Vendor I |  |  |  | Vendor II |  |  |  |
| --- | --- | --- | --- | --- | --- | --- | --- | --- | --- |
|  |  | Non-ambient<br>(mean [sd]) | Ambient<br>(mean [sd]) | Mean<br>difference<br>(95% CI)<sup>a</sup> | P-value | Non-ambient<br>(mean [sd]) | Ambient<br>(mean [sd]) | Mean<br>difference<br>(95% CI)<sup>a</sup> | P-value |
| Mean Sentence Length (# of word tokens) | HPI | 16.61 (12.68) | 17.96 (6.43) | 1.345 (-0.662, 3.352) | 0.186 | 17.86 (18.21) | 18.46 (16.35) | 0.596 (-0.997, 2.189) | 0.462 |
|  | A&P | 25.81 (24.25) | 20.52 (22.45) | -5.365 (-12.477, 1.747) | 0.137 | 22.77 (20.24) | 22.60 (24.68) | -0.173 (-3.939, 3.593) | 0.928 |
| Mean Clause Length (# of word tokens of T-unit) | HPI | 12.69 (5.02) | 14.12 (2.50) | 1.434 (0.748, 2.121) | <0.001 | 13.39 (6.92) | 14.14 (5.05) | 0.753 (0.045, 1.460) | 0.037 |
|  | A&P | 18.35 (9.69) | 15.34 (6.64) | -3.038 (-5.963, -0.114) | 0.042 | 16.97 (8.85) | 16.56 (8.80) | -0.417 (-1.683, 0.849) | 0.517 |
| Dependency Distance | HPI | 2.84 (0.55) | 3.00 (0.30) | 0.154 (0.102, 0.207) | <0.001 | 2.88 (0.62) | 2.94 (0.47) | 0.052 (-0.007, 0.112) | 0.086 |
|  | A&P | 3.34 (0.83) | 3.09 (0.74) | -0.257 (-0.477, -0.036) | 0.023 | 3.19 (0.82) | 3.13 (0.74) | -0.066 (-0.167, 0.035) | 0.201 |

HPI: history of present illness; A&P: assessment and plan
<sup>a</sup> Mean differences were estimated using patient-level pre–post differences by fitting intercept-only linear models within each ambient listening system. Robust standard errors were clustered at the provider level to account for potential correlation in outcomes among patients treated by the same provider.

Figure 2 presents the comparison of POS distributions and POS entropy between ambient and non-ambient documentation in the HPI and A&P sections for Vendor I and Vendor II. Panels a) and c) display the proportion of POS tags in the HPI section for Vendor I and Vendor II, respectively, while panels b) and d) present the corresponding POS entropy comparisons. Panels e) and g) illustrate the proportion of POS tags in the A&P section for Vendor I and Vendor II, respectively, and panels f) and h) show the POS entropy comparisons for the A&P section.

**Figure 2.**
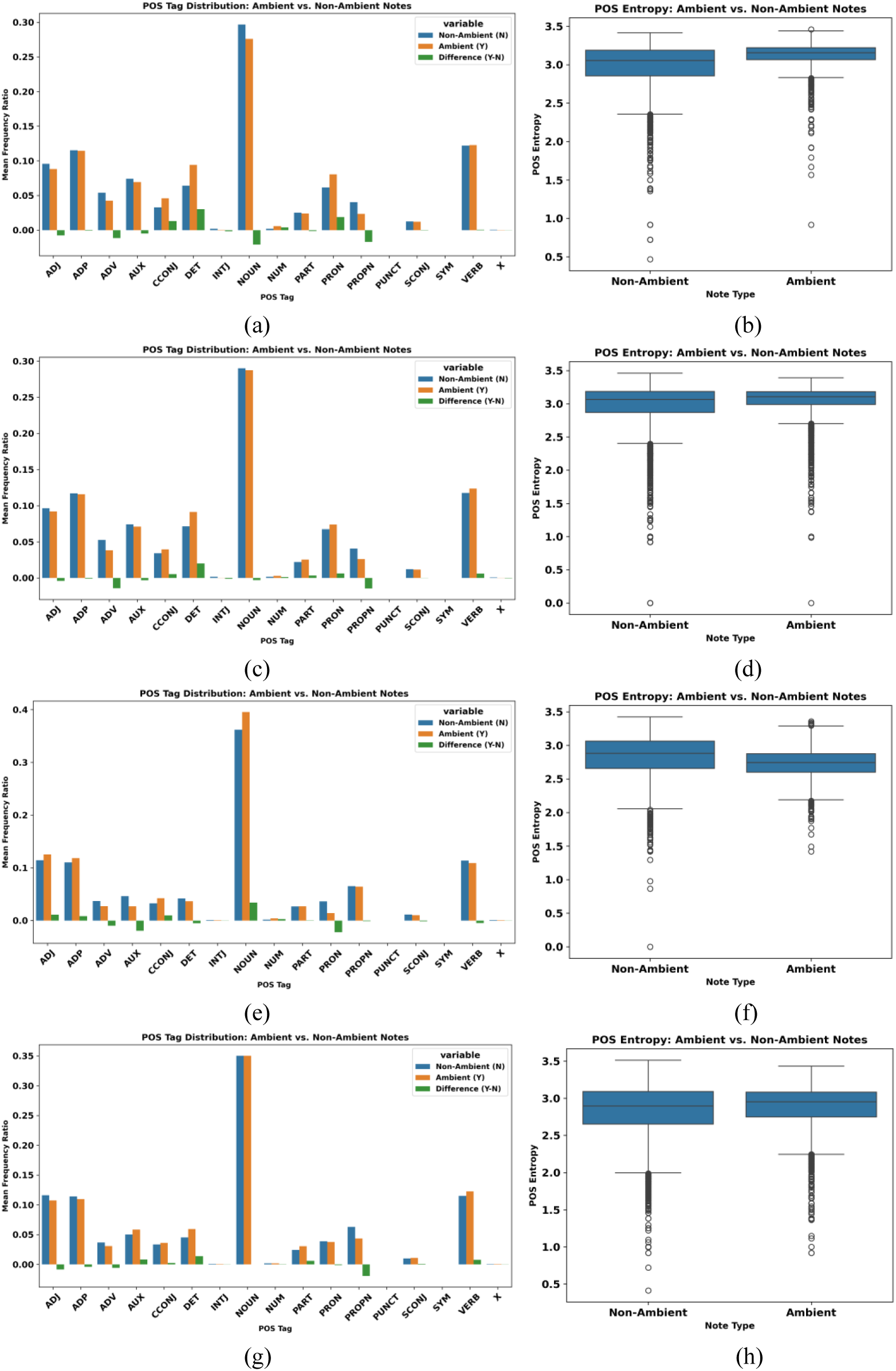
Proportion of POS tag between ambient and non-ambient in HPI section for a) Vendor I c) Vendor II POS entropy between ambient and non-ambient in HPI section for b) Vendor I d) Vendor II Proportion of POS tag between ambient and non-ambient in A&P section for e) Vendor I g) Vendor II POS entropy between ambient and non-ambient in A&P section for f) Vendor I h) Vendor II

In the HPI section, panels a) and b) demonstrate that Vendor I’s ambient notes exhibit a marked increase in linguistic diversity. POS entropy is substantially higher in ambient documentation (Δ = 0.158, *P* < 0.001). This increase is accompanied by higher proportions of determiners, pronouns, coordinating conjunctions, and numerals, with determiners showing the largest shift (Δ = 0.030, *P* < 0.001). In contrast, nouns, proper nouns, and adverbs decrease in relative frequency. The reduction in noun and proper noun density suggests a shift away from telegraphic, content-heavy phrasing toward more syntactically elaborated narrative structure.

Panels c) and d) show a similar but more moderate pattern for Vendor II in the HPI section. POS entropy increases in ambient notes (Δ = 0.080, *P* < 0.001), though the magnitude is approximately half that observed for Vendor I. Ambient notes demonstrate increased use of determiners, verbs, conjunctions, and particles, alongside reductions in adverbs and proper nouns. While both vendors exhibit increased syntactic diversity in HPI documentation, the effect is more pronounced for Vendor I.

In the A&P section, panels e) and f) reveal a contrasting pattern for Vendor I. Ambient notes demonstrate a decrease in POS entropy (Δ = −0.090, *P* < 0.001), indicating reduced linguistic variability. At the same time, ambient A&P documentation shows higher proportions of nouns, adjectives, and coordinating conjunctions, along with marked reductions in pronouns and auxiliary verbs. The increase in noun density (Δ = 0.034, *P* < 0.001) combined with reduced pronoun and auxiliary usage suggests a shift toward more content-dense and structurally consolidated documentation in the assessment and plan.

In contrast, panels g) and h) indicate that Vendor II’s ambient A&P notes exhibit a modest increase in POS entropy (Δ = 0.053, *P* < 0.001). Ambient documentation shows increased use of determiners, auxiliaries, verbs, and particles, with a notable reduction in proper nouns. Unlike Vendor I, Vendor II does not demonstrate a strong shift toward noun-dominant compression in the A&P section and instead maintains or slightly increases syntactic variability.

Overall, Figure 2 illustrates that the linguistic impact of ambient documentation is both vendor-dependent and section-specific. In the HPI section (panels a–d), both vendors increase syntactic diversity, with Vendor I showing a stronger expansion effect. In the A&P section (panels e–h), Vendor I shifts toward a more structured and noun-heavy style with reduced entropy, whereas Vendor II maintains or modestly increases linguistic variability.

Figure 3 presents adjusted marginal mean patient-level pre–post differences in mean sentence length and mean clause length for each level of provider sex, specialty, and type across both the HPI and A&P sections, further stratified by vendor system.

**Figure 3.**
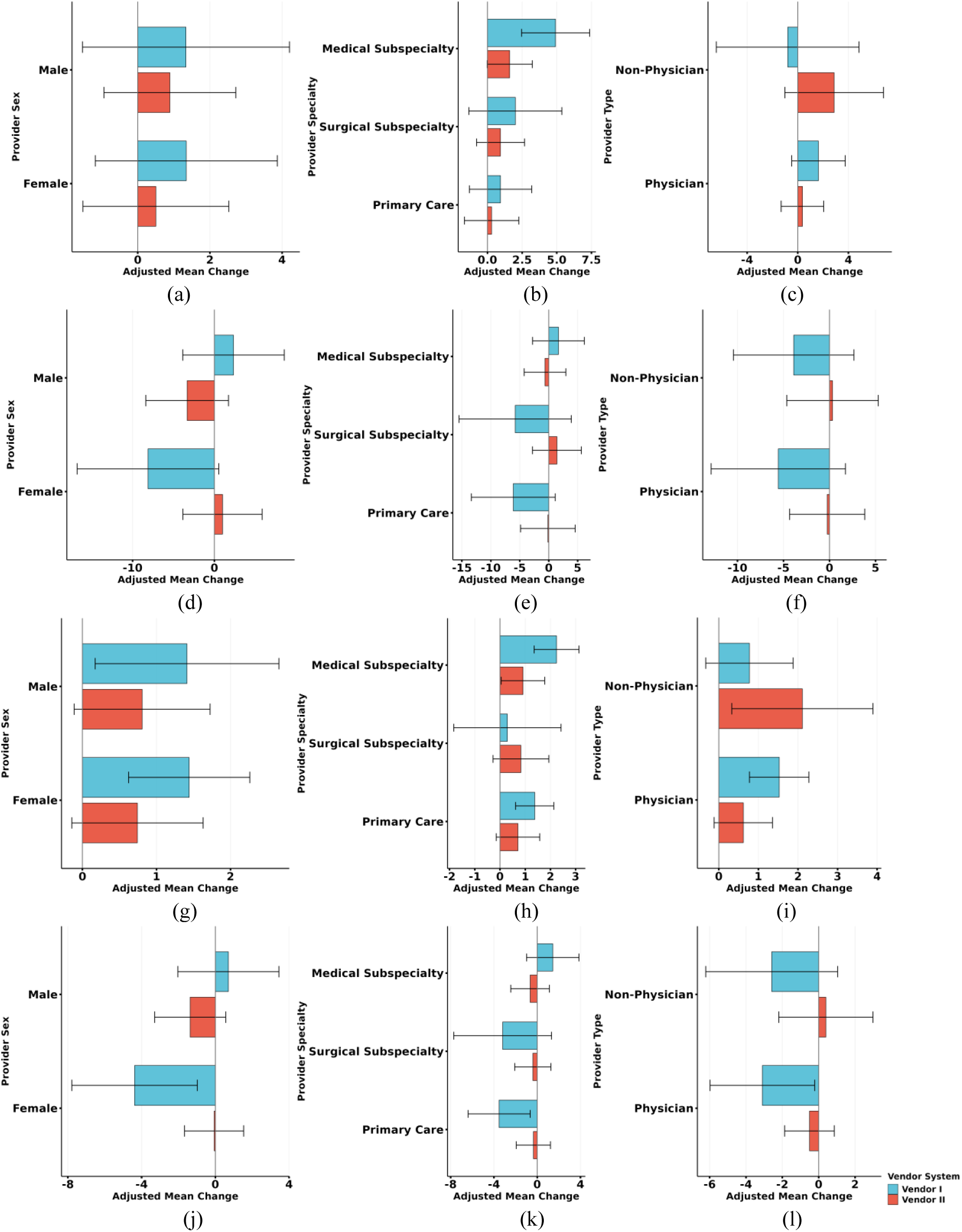
Mean sentence length for HPI section in (a) Sex (b) Provider Specialty (c) Provider Type Mean sentence length for A&P section in (d) Sex (e) Provider Specialty (f) Provider Type Name Mean clause length for HPI section in (g) Sex (h) Provider Specialty (i) Provider Type Mean clause length for A&P section in (j) Sex (k) Provider Specialty (l) Provider Type

Panels (a), (b), and (c) show mean sentence length changes in the HPI section stratified by provider sex, specialty, and provider type, respectively. In panel (a), Vendor I demonstrated small, non-significant increases for female (1.35 words, 95% CI: −1.17 to 3.87, *P* = 0.291) and male providers (1.34 words, 95% CI: −1.53 to 4.20, *P* = 0.356). Vendor II showed similarly small, non-significant changes for females (0.50 words, *P* = 0.624) and males (0.89 words, *P* = 0.336). In panel (b), stratified by provider specialty, Vendor I showed a significant increase among medical subspecialties (4.91 words, 95% CI: 2.46 to 7.37, *P* < 0.001), whereas primary care and surgical subspecialties showed non-significant changes. Vendor II demonstrated smaller, non-significant increases, with a borderline effect for medical subspecialties (1.61 words, 95% CI: −0.02 to 3.23, *P* = 0.053). In panel (c), neither vendor demonstrated statistically significant changes in HPI sentence length by provider type.

Panels (d), (e), and (f) present mean sentence length changes in the A&P section stratified by provider sex, specialty, and provider type, respectively. In panel (d), Vendor I showed a notable but non-significant decrease among female providers (−8.12 words, 95% CI: −16.78 to 0.54, *P* = 0.066), while Vendor II showed minimal changes for both sexes. In panel (e), Vendor I showed decreases among primary care (−6.12 words, *P* = 0.096) and surgical subspecialties (−5.79 words, *P* = 0.238), though none reached statistical significance; Vendor II demonstrated negligible changes across specialties. In panel (f), neither vendor showed significant sentence length changes by provider type in the A&P section.

Panels (g), (h), and (i) display mean clause length changes in the HPI section stratified by provider sex, specialty, and provider type, respectively. In panel (g), Vendor I demonstrated significant increases for female (1.44 words, 95% CI: 0.62 to 2.26, *P* = 0.001) and male providers (1.41 words, 95% CI: 0.17 to 2.65, *P* = 0.026), while Vendor II showed smaller, non-significant increases. In panel (h), Vendor I showed significant increases among primary care (1.37 words, *P* = 0.001) and medical subspecialties (2.24 words, *P* < 0.001), whereas Vendor II demonstrated a significant increase only among medical subspecialties (0.91 words, 95% CI: 0.05 to 1.77, *P* = 0.039). In panel (i), Vendor I showed a significant increase among physicians (1.52 words, *P* < 0.001), while Vendor II demonstrated a significant increase among non-physicians (2.11 words, 95% CI: 0.33 to 3.90, *P* = 0.021).

Panels (j), (k), and (l) show mean clause length changes in the A&P section stratified by provider sex, specialty, and provider type, respectively. In panel (j), Vendor I demonstrated a significant decrease among female providers (−4.38 words, 95% CI: −7.78 to −0.97, *P* = 0.012), whereas Vendor II showed no significant changes. In panel (k), Vendor I showed a significant reduction among Primary Care providers (−3.52 words, 95% CI: −6.39 to −0.65, *P* = 0.017), with no significant findings for Vendor II. In panel (l), Vendor I demonstrated a significant decrease among Physicians (−3.10 words, 95% CI: −5.98 to −0.22, *P* = 0.035), while Vendor II again showed no significant changes.

Overall, Figure 3 indicates that structural changes were more pronounced in the HPI section than in the A&P section, and effects were generally stronger and more consistent for Vendor I than for Vendor II, particularly for clause-level complexity.

Supplement Figure 1 shows adjusted patient-level pre–post changes in dependency distance for the HPI section and (panels a–c) A&P section (panels d–f), further stratified by vendor system. In the HPI section, Vendor I showed consistent increases across most groups, including both sexes, primary care, medical subspecialties, and physicians (*P* < 0.001). Vendor II showed smaller increases, reaching significance for male providers only (*P* = 0.021). In the A&P section, Vendor I demonstrated significant decreases among female providers, primary care, and physicians (*P* < 0.05), while Vendor II showed a significant decrease only among male providers (*P* = 0.042). Overall, HPI dependency distance increased (especially Vendor I), whereas A&P changes were modest and selective.

Table 2 presents ambient versus non-ambient comparisons for lexical ambiguity, stratified by vendor and note section. Across all licenses, ambient documentation was associated with lower lexical ambiguity, particularly in HPI sections. For example, HPI lexical ambiguity decreased under Vendor I (6.65 to 5.81, mean difference −0.838, *P*<0.001), and Vendor II (6.58 to 5.72, *P*<0.001). A&P sections also showed reductions, though effect sizes were smaller: Vendor I (5.68 to 5.27, *P*<0.001), and Vendor II (5.78 to 5.55, *P*=0.002).

**Table 2.** Ambient vs. non-ambient comparisons across vendors by note section type, with lexical ambiguity.

|  | Vendor I |  |  |  | Vendor II |  |  |  |
| --- | --- | --- | --- | --- | --- | --- | --- | --- |
|  | Non-ambient (mean [sd]) | Ambient (mean [sd]) | Mean difference (95% CI)<sup>a</sup> | P-value | Non-ambient (mean [sd]) | Ambient (mean [sd]) | Mean difference (95% CI)<sup>a</sup> | P-value |
| **HPI** | 6.65 (1.37) | 5.81 (0.81) | -0.838 (-1.019, -0.656) | <0.001 | 6.58 (1.34) | 5.72 (0.77) | -0.859 (-1.026, -0.692) | <0.001 |
| **A&P** | 5.68 (0.98) | 5.27 (0.70) | -0.413 (-0.605, -0.221) | <0.001 | 5.78 (0.97) | 5.55 (0.70) | -0.233 (-0.381, -0.085) | 0.002 |

**HPI:** history of present illness; **A&P:** assessment and plan
<sup>a</sup> Mean differences were estimated using patient-level pre–post differences by fitting intercept-only linear models within each ambient listening system. Robust standard errors were clustered at the provider level to account for potential correlation in outcomes among patients treated by the same provider.

Figure 4 presents adjusted marginal changes for each level of provider sex, specialty, and type in lexical ambiguity in the HPI (a–c) and A&P (d–f) sections, further stratified by vendor system. In the HPI section, both Vendor I and Vendor II demonstrated substantial and statistically significant reductions in lexical ambiguity for female and male providers (all *P* ≤ 0.001). Reductions were also significant for primary care and medical subspecialties for both vendors (all *P* ≤ 0.001), whereas surgical subspecialties showed no significant change. By provider type, physicians in both vendors exhibited significant decreases (both *P* < 0.001), while non-physicians showed a nonsignificant reduction for Vendor I (*P* = 0.094) but a significant decrease for Vendor II (*P* = 0.001).

**Figure 4.**
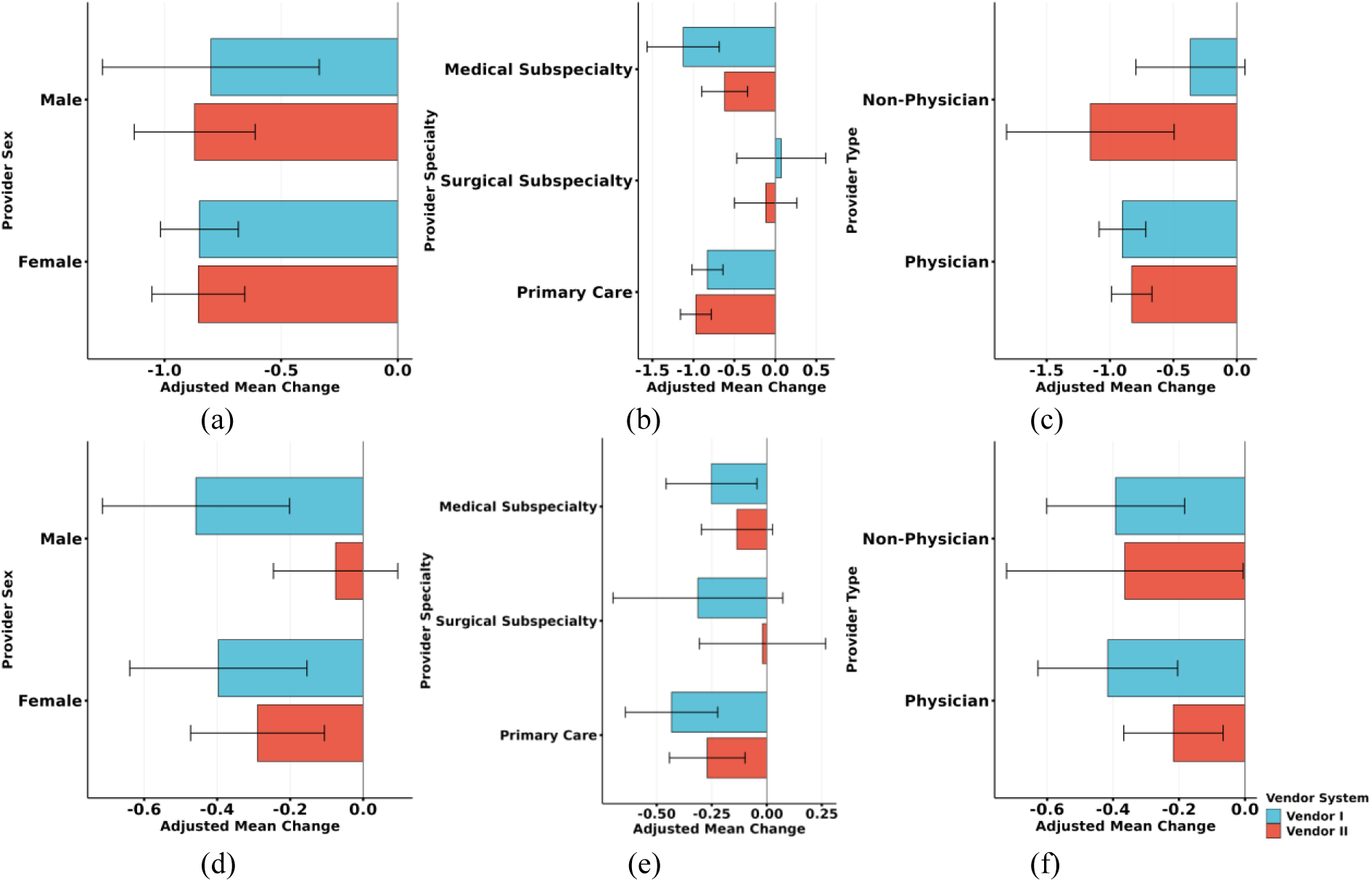
Lexical ambiguity for HPI section in (a) Sex (b) Provider Specialty (c) Provider type. Lexical ambiguity for A&P section in (d) Sex (e) Provider Specialty (f) Provider type.

In the A&P section, reductions in lexical ambiguity were smaller in magnitude. For Vendor I, decreases were significant across sex, primary care, medical subspecialty, and both provider types (all *P* ≤ 0.018), except for surgical subspecialties (*P* = 0.110). For Vendor II, significant reductions were observed for female providers, primary care, physicians, and non-physicians (*P* = 0.002–0.047), whereas changes for males, surgical subspecialties, and medical subspecialties were not statistically significant. Overall, reductions were more consistent and pronounced in the HPI than in the A&P section across both vendors.

Table 3 presents ambient versus non-ambient comparisons for linguistic variability, stratified by vendor and note section. Ambient documentation consistently showed substantial reductions across all licenses and both models, particularly in A&P section. For example, A&P decreased from 214.28 to 152.02 for Vendor I (mean difference −62.93, *P*<0.001) using BioGPT, with similar patterns observed for GPT-2 (e.g., Vendor I A&P: 74.71 to 47.57, *P*<0.001). In contrast, results for HPI alone were more heterogeneous: GPT-2 demonstrated significant ambient-related reductions across all licenses (e.g., Vendor II: 176.41 to 54.55, *P*<0.001), whereas BioGPT estimates for HPI exhibited large variability and were not statistically significant for Vendor II.

**Table 3.**
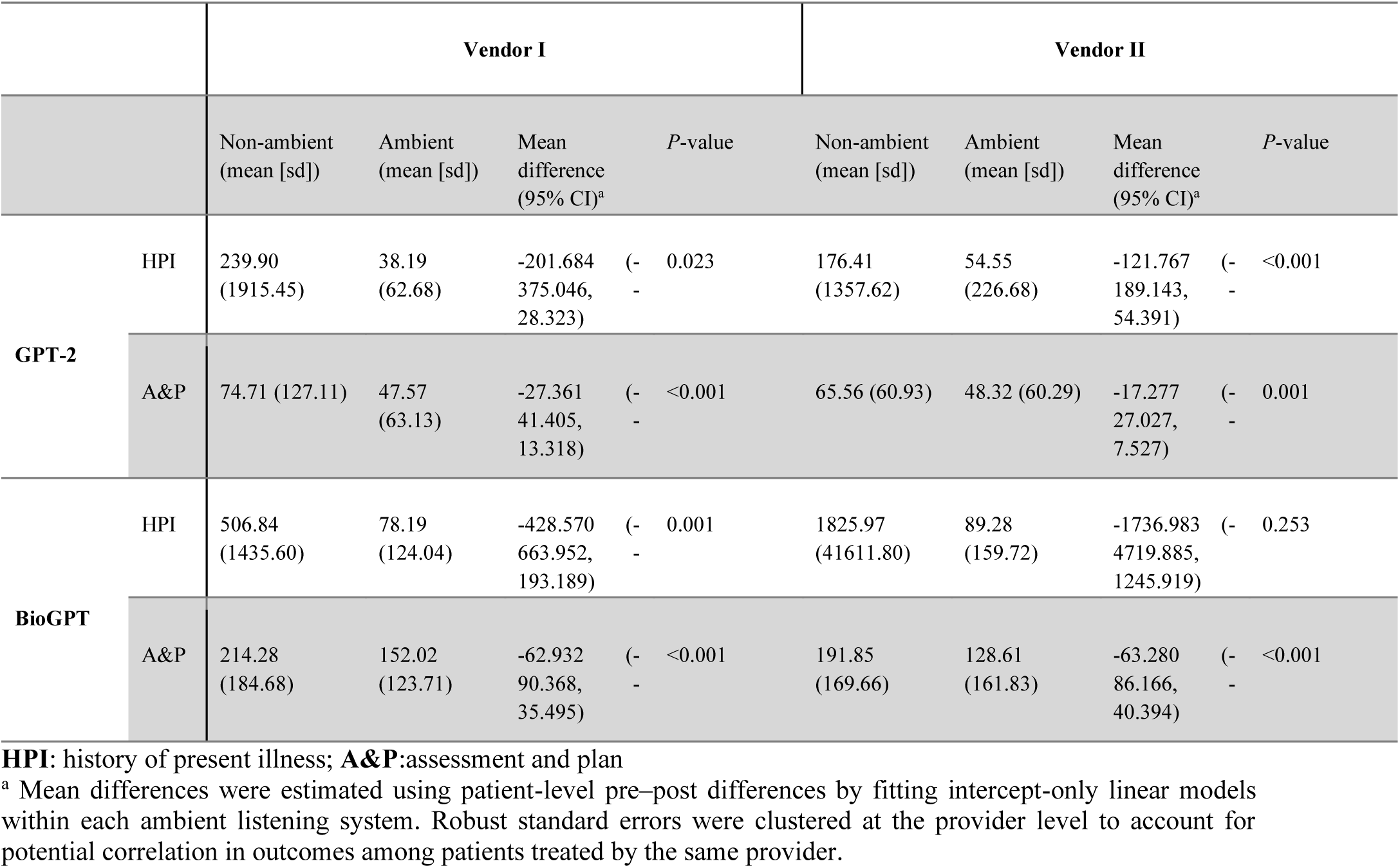
Ambient vs. non-ambient comparisons across vendors by note section type, with linguistic variability.

Figure 5 illustrates adjusted marginal changes in perplexity for each level of provider sex, specialty, and type, a measure of linguistic variability, across HPI and A&P sections for GPT-2 (panels a–f) and BioGPT (panels g–l), further stratified by vendor system. For the HPI section (panels a–c), GPT-2 generally reduced perplexity, indicating less variability and more predictable language. Vendor I showed significant reductions for female providers (*P* < 0.001), primary care providers (*P* = 0.016), and physicians (*P* = 0.017), whereas reductions were nonsignificant for males, and medical subspecialty providers. Vendor II demonstrated smaller but still significant decreases for female providers (*P* = 0.001), primary care (*P* = 0.004), and physicians (*P* < 0.001).

**Figure 5.**
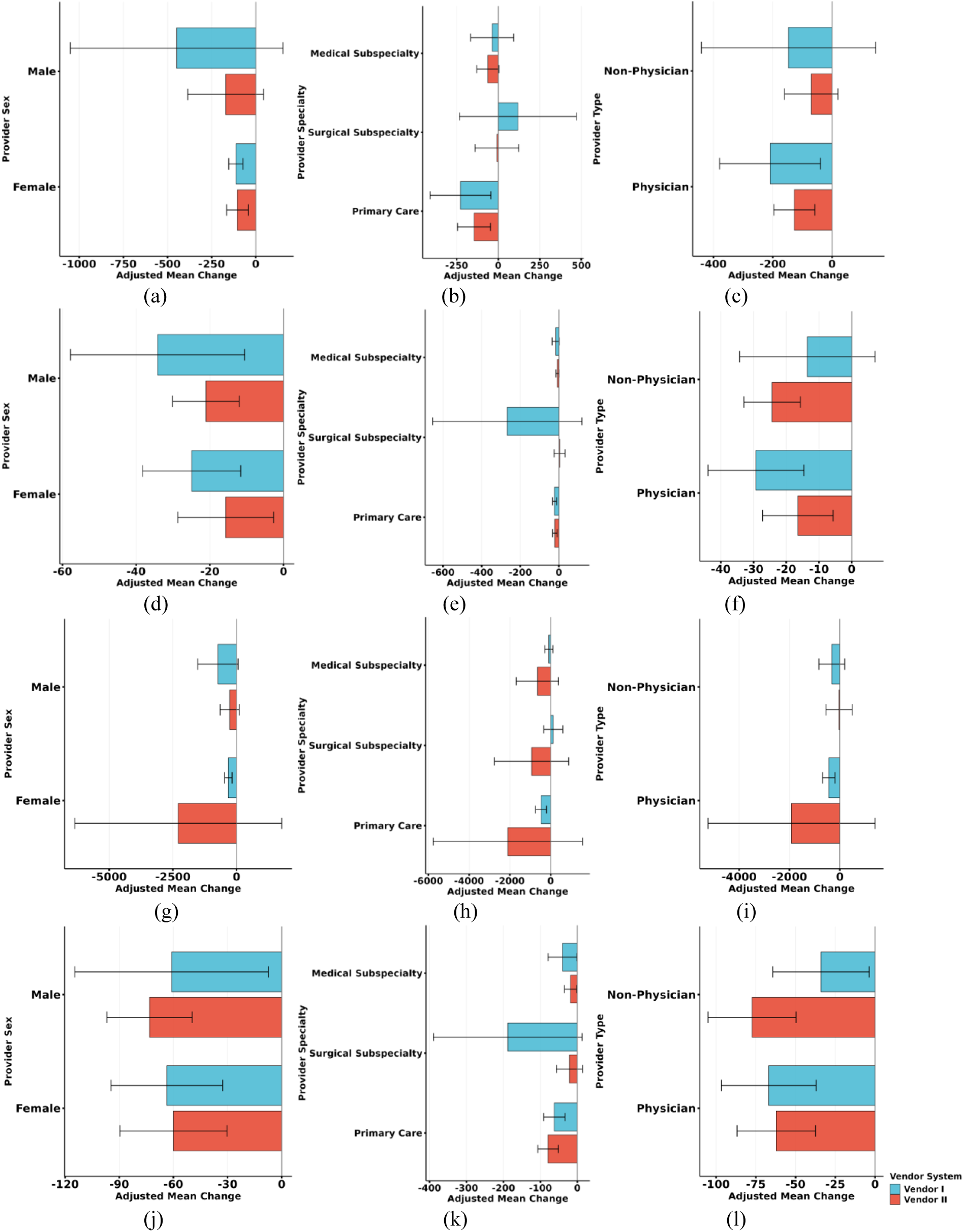
Linguistic variability by GPT-2 for HPI section in (a) Sex (b) Provider Specialty (c) Provider Type Linguistic variability by GPT-2 for A&P section in (d) Sex (e) Provider Specialty (f) Provider Type Linguistic variability by BioGPT for HPI section in (g) Sex (h) Provider Specialty (i) Provider Type Linguistic variability by BioGPT for A&P section in (j) Sex (k) Provider Specialty (l) Provider Type

In the A&P section (panels d–f), GPT-2 reduced perplexity across most subgroups, with Vendor I showing consistent significant reductions for female, male, primary care, and physicians (*P* ≤ 0.005), while Vendor II had modest but significant decreases for most groups except medical subspecialties. Non-physician providers showed variable responses, with significance only in Vendor II (*P* < 0.001 for male non-physicians).

For BioGPT, the HPI section (panels g–i) demonstrated reductions in perplexity for Vendor I (except for surgical subspecialty), particularly among females (*P* < 0.001), primary care (*P* = 0.001), and physicians (*P* = 0.001), reflecting more standardized language generation. Vendor II showed inconsistent reductions with large SEs and mostly nonsignificant differences, suggesting higher variability or less consistent improvement across providers.

In the A&P section (panels j–l), BioGPT reduced perplexity for most groups in Vendor I, with significant decreases for female, male, primary care, medical subspecialty, physicians, and non-physicians (*P* ≤ 0.044). Vendor II also showed significant reductions across most subgroups, including females, males, primary care, medical subspecialty, physicians, and non-physicians (*P* ≤ 0.028), although effects were slightly smaller than Vendor I.

Overall, GPT-2 and BioGPT both decreased perplexity, indicating reduced linguistic variability and more consistent documentation. Vendor I typically exhibited larger and more consistent reductions across provider sex, specialty, and type, whereas Vendor II showed smaller or inconsistent effects, particularly for HPI with BioGPT. These results highlight differences in model performance and language standardization across vendors and documentation sections.

Table 4 summarizes ambient versus non-ambient differences in local and global coherence, shuffled local and global coherence, and the shuffled difference across vendors for HPI and A&P sections. Across all licenses, ambient documentation was consistently associated with higher local coherence in HPI, with increases observed for Vendor I (0.84 to 0.87, *P*<0.001), and Vendor II (0.84 to 0.86, *P*<0.001). Similar improvements in local coherence were observed for A&P under Vendor I, whereas effects for Vendor II were attenuated and not statistically significant after adjustment in A&P sections.

**Table 4.**
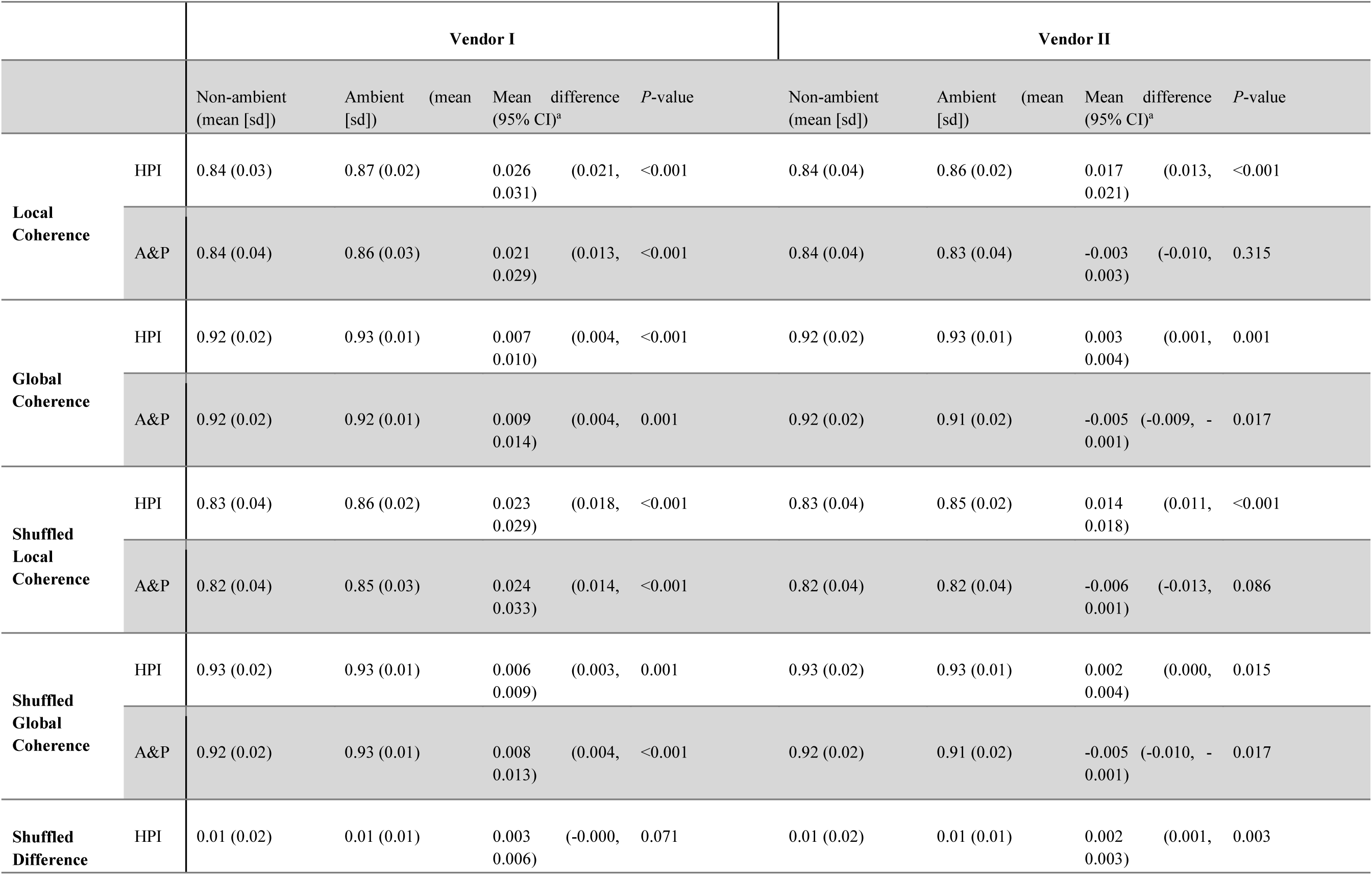
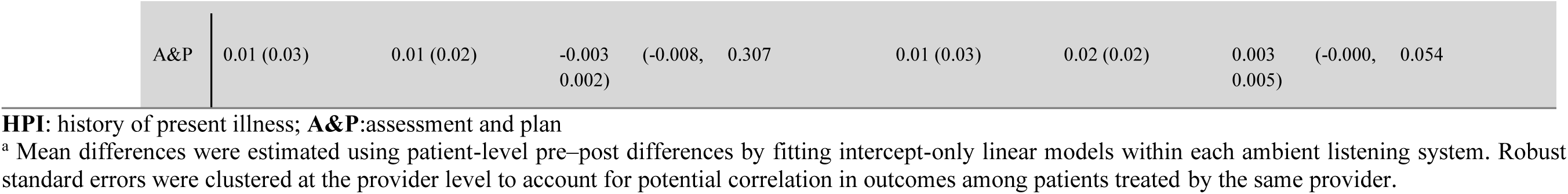
Ambient vs. non-ambient comparisons across vendors by note section type, with local coherence, global coherence, shuffled local coherence, shuffled global coherence, and shuffled difference.

For global coherence, ambient notes demonstrated modest but statistically significant increases in HPI across all licenses, including Vendor I (0.92 to 0.93, *P*<0.001), and Vendor II (0.92 to 0.93, *P*=0.001). In contrast, A&P sections showed small decreases or null effects in global coherence for Vendor II (−0.005, *P*=0.017), while Vendor I exhibited a modest increase (*P*=0.001).

Across all licenses, ambient documentation was associated with higher shuffled local coherence in HPI, with consistent increases for Vendor I (0.83 to 0.86, *P*<0.001), and Vendor II (0.83 to 0.85, *P*<0.001). Similar increases were observed for A&P under Vendor I (0.82 to 0.85, *P*<0.001), whereas effects for Vendor II in A&P were small and not statistically significant after adjustment.

For shuffled global coherence, ambient notes demonstrated small but statistically significant increases in HPI across all vendors, including Vendor I (mean difference 0.006, *P*=0.001), and Vendor II (*P*=0.015). In A&P sections, shuffled global coherence decreased slightly under Vendor II (−0.005, *P*=0.017), while remaining stable or modestly increased under Vendor I (0.92 to 0.93, *P*<0.001).

Figure 6 illustrates the adjusted marginal changes for each level of provider sex, specialty, and type in local and global coherence for clinical documentation across the HPI and A&P sections, further stratified by vendor system. In the HPI section, local coherence increased significantly for most subgroups in Vendor I, including female and male providers, primary care providers, physicians, and non-physicians, with *P*-values <0.001. Surgical subspecialty providers showed nonsignificant changes (*P* = 0.093), while medical subspecialty providers had modest but significant improvements (*P* = 0.007). Vendor II exhibited smaller increases overall, with significant improvements in female and male providers, primary care, medical subspecialty, and provider type (*P* ≤0.001), whereas surgical subspecialty remained nonsignificant (*P* = 0.789). Global coherence in the HPI section showed modest but significant increases in Vendor I for most subgroups, including female and male providers, primary care, medical subspecialty, and physicians (*P* ≤0.009), with surgical subspecialty remaining nonsignificant (*P* = 0.279). Vendor II showed smaller improvements, with some subgroups, such as surgical subspecialty and physician type, exhibiting marginal significance (*P* = 0.005–0.671).

**Figure 6.**
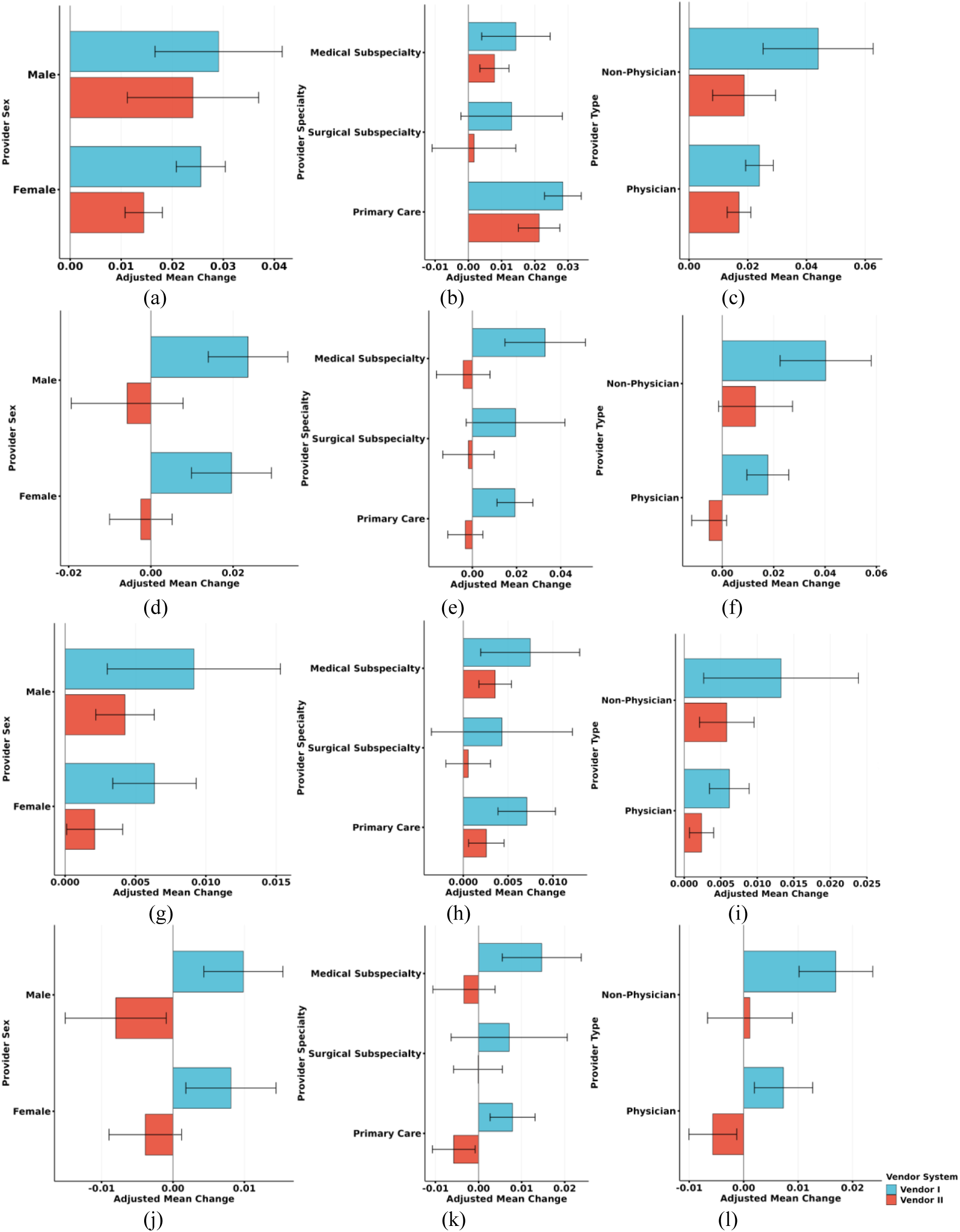
Local coherence for HPI section in (a) Sex (b) Provider Specialty (c) Provider Type. Local coherence for A&P section in (d) Sex (e) Provider Specialty (f) Provider Type. Global coherence for HPI section in (g) Sex (h) Provider Specialty (i) Provider Type Global coherence for A&P section in (j) Sex (k) Provider Specialty (l) Provider Type

In the A&P section, local coherence increased significantly in Vendor I for female and male providers, primary care, medical subspecialty, physicians, and non-physicians (*P* ≤0.001), while surgical subspecialty providers showed nonsignificant changes (*P* = 0.086). Vendor II generally showed minimal changes in local coherence, with none reaching significance except for a slight trend in non-physicians (*P* = 0.078). Global coherence in the A&P section increased modestly for most subgroups in Vendor I (*P* = 0.001–0.012), with surgical subspecialty remaining nonsignificant (*P* = 0.296). Vendor II exhibited small decreases or negligible changes across several subgroups, including male providers, primary care, and physician type, some of which were statistically significant (*P* = 0.012–0.027), suggesting slight variability or reduction in global coherence in these cases.

Overall, Vendor I consistently improved both local and global coherence across HPI and A&P sections, with the largest effects observed for provider sex, primary care specialty, and provider type. In contrast, Vendor II showed smaller and less consistent changes, with some subgroups demonstrating nonsignificant or slightly negative differences, particularly in the A&P section, indicating greater variability in documentation coherence.

Supplement Figure 2 presents the adjusted marginal changes for each level of provider sex, specialty, and type in shuffled local and global coherence, as well as shuffled differences, across the HPI and A&P sections, further stratified by vendor system. Shuffled local coherence showed consistent positive changes in Vendor I across nearly all subgroups in both HPI and A&P sections, with the largest increases observed for non-physician providers (*P* ≤0.001), while Vendor II exhibited smaller and less consistent changes, with several nonsignificant results, particularly in the A&P section (*P* = 0.106–0.380). Shuffled global coherence followed a similar pattern, with Vendor I showing modest but significant improvements across most subgroups (*P* = 0.001–0.031), whereas Vendor II showed minimal changes, with some small decreases reaching significance in male providers (*P* = 0.002–0.023). The shuffled differences, reflecting the net change after accounting for randomization, were generally close to zero for most subgroups across both sections and vendors, indicating limited overall effect, though some small but statistically significant changes were observed for certain subgroups in Vendor I and Vendor II (*P* = 0.001–0.046). Overall, the figure suggests that while Vendor I consistently improved shuffled coherence measures, Vendor II showed more variable and often negligible changes, highlighting differences in documentation coherence stability across vendors.

Table 5 presents ambient versus non-ambient comparisons for word count, complex word count, sentence count, medical terminology usage and readability stratified by vendor and note section. Across all licenses, ambient documentation was associated with substantially higher word counts in HPI, with consistent increases for Vendor I (128.1 to 225.5, mean difference 97.43, *P*<0.001), and Vendor II (156.0 to 245.5, *P*<0.001). A&P sections showed smaller increases, reaching statistical significance primarily under Vendor II.

**Table 5.** Ambient vs. non-ambient comparisons across vendors by note section type, with word count, complex word count, sentence count, medical term density and MedReadMe Index.

|  |  | Vendor I |  |  |  |  | Vendor II |  |  |  |
| --- | --- | --- | --- | --- | --- | --- | --- | --- | --- | --- |
|  |  | Non-ambient (mean [sd]) | Ambient (mean [sd]) | Mean difference (95% CI)<sup>a</sup> | P-value |  | Non-ambient (mean [sd]) | Ambient (mean [sd]) | Mean difference (95% CI)<sup>a</sup> | P-value |
| Word Count | HPI | 128.10 (126.22) | 225.53 (138.53) | 97.428 (69.967, 124.890) | <0.001 |  | 155.97 (159.45) | 245.50 (155.14) | 89.529 (68.732, 110.325) | <0.001 |
|  | A&P | 220.00 (183.30) | 237.15 (144.65) | 17.152 (-18.255, 52.559) | 0.338 |  | 226.45 (189.98) | 257.37 (192.44) | 30.920 (12.769, 49.072) | 0.001 |
| Complex Word Count | HPI | 31.31 (30.20) | 61.67 (30.61) | 30.363 (23.731, 36.994) | <0.001 |  | 36.58 (35.70) | 71.09 (38.35) | 34.503 (29.077, 39.928) | <0.001 |
|  | A&P | 64.48 (45.43) | 82.20 (42.39) | 17.720 (8.748, 26.692) | <0.001 |  | 64.49 (43.87) | 78.55 (48.05) | 14.068 (9.769, 18.366) | <0.001 |
| Sentence Count | HPI | 11.16 (11.71) | 15.43 (12.49) | 4.272 (1.810, 6.734) | 0.001 |  | 12.76 (12.17) | 17.85 (11.50) | 5.087 (3.155, 7.018) | <0.001 |
|  | A&P | 15.95 (15.83) | 19.22 (10.63) | 3.273 (-0.971, 7.518) | 0.129 |  | 16.14 (13.86) | 17.34 (12.42) | 1.202 (-0.800, 3.203) | 0.238 |
| Medical Term Density | HPI | 0.56 (0.24) | 0.57 (0.16) | 0.011 (-0.019, 0.041) | 0.463 |  | 0.56 (0.22) | 0.58 (0.18) | 0.023 (-0.001, 0.047) | 0.056 |
|  | A&P | 0.58 (0.19) | 0.68 (0.16) | 0.095 (0.067, 0.123) | <0.001 |  | 0.59 (0.17) | 0.62 (0.17) | 0.030 (0.017, 0.042) | <0.001 |
| MedReadMe | HPI | 3.70 (0.39) | 3.81 (0.23) | 0.113 (0.047, 0.180) | 0.001 |  | 3.71 (0.40) | 3.81 (0.26) | 0.097 (0.046, 0.147) | <0.001 |
|  | A&P | 3.95 (0.39) | 3.96 (0.25) | 0.012 (-0.090, 0.114) | 0.814 |  | 3.90 (0.37) | 3.88 (0.34) | -0.026 (-0.076, 0.023) | 0.298 |

**HPI:** history of present illness; **A&P:**assessment and plan <sup>a</sup> Mean differences were estimated using patient-level pre–post differences by fitting intercept-only linear models within each ambient listening system. Robust standard errors were clustered at the provider level to account for potential correlation in outcomes among patients treated by the same provider.

For complex word count, ambient notes consistently contained more complex words across HPI, including Vendor I (31.3 to 61.7, mean difference 30.36, *P*<0.001), and Vendor II (36.6 to 71.1, *P*<0.001). A&P section also demonstrated increases, though effect sizes were smaller and varied by vendors.

Analysis of sentence count revealed moderate increases under ambient conditions, particularly in HPI section across all vendors, with mean differences ranging from ∼4 to 5 sentences (all *P*≤0.001). In A&P section, sentence count changes were smaller and less consistent, occasionally not reaching statistical significance.

Across vendors, Ambient documentation was generally associated with higher Medical Term Density, particularly in the A&P section. For Vendor I, Ambient A&P notes exhibited a marked increase in medical term density from 0.58 (SD 0.19) to 0.68 (SD 0.16), yielding a mean difference of 0.095 (95% CI: 0.067 to 0.123; *P* < 0.001). Vendor II demonstrated similar patterns, with significant increases in A&P (mean difference: 0.030; 95% CI: 0.017 to 0.042; *P* < 0.001). Across all vendors, Ambient conditions are consistently associated with significantly improved readability in the HPI section. For Vendor I, Ambient HPI readability increases from 3.70 (SD 0.39) to 3.81 (SD 0.23), yielding a mean difference of 0.113 (95% CI: 0.047 to 0.180; *P* = 0.001). A similar pattern is observed for Vendor II, where HPI readability improves from 3.71 (SD 0.40) to 3.81 (SD 0.26), with a mean difference of 0.097 (95% CI: 0.046 to 0.147; *P* < 0.001). In contrast, A&P readability shows no statistically significant differences between Ambient and Non-Ambient conditions for any vendor, and effect sizes are small and directionally inconsistent.

Supplement Figure 3 summarizes the adjusted marginal changes for each level of provider sex, specialty, and type in word count, complicated word count, and sentence count across the HPI and A&P sections, further stratified by vendor system. For the HPI section, Vendor I consistently showed significant increases in word count across all subgroups, with the largest increases observed for non-physician providers (*P* < 0.001), while Vendor II also showed significant increases but with slightly smaller magnitudes. In the A&P section, changes in word count were more variable, with Vendor I showing mostly nonsignificant changes except for surgical subspecialties (*P* = 0.019), whereas Vendor II demonstrated modest but significant increases in several subgroups, particularly males, primary care, and non-physicians (*P* ≤0.042). Complicated word counts increased significantly across most subgroups for both vendors in the HPI section, reflecting more complex language use, while in the A&P section, increases were generally smaller and less consistent, especially for non-physician providers in Vendor I (*P* = 0.398). Sentence counts followed a similar pattern, with Vendor I showing significant increases in HPI sentences for most subgroups (*P* ≤0.039) and smaller or nonsignificant changes in the A&P section, whereas Vendor II showed modest increases in HPI sentences and mostly nonsignificant changes in the A&P section, with a few exceptions such as males (*P* = 0.045). Overall, the figure indicates that both vendors contributed to increases in text length and complexity, particularly in the HPI section, but these effects were less pronounced and more variable in the A&P section.

Figure 7 illustrates adjusted marginal changes for each level of provider sex, specialty, and type in medical term density and MedReadMe scores across the HPI and A&P sections, further stratified by vendor system.

**Figure 7.**
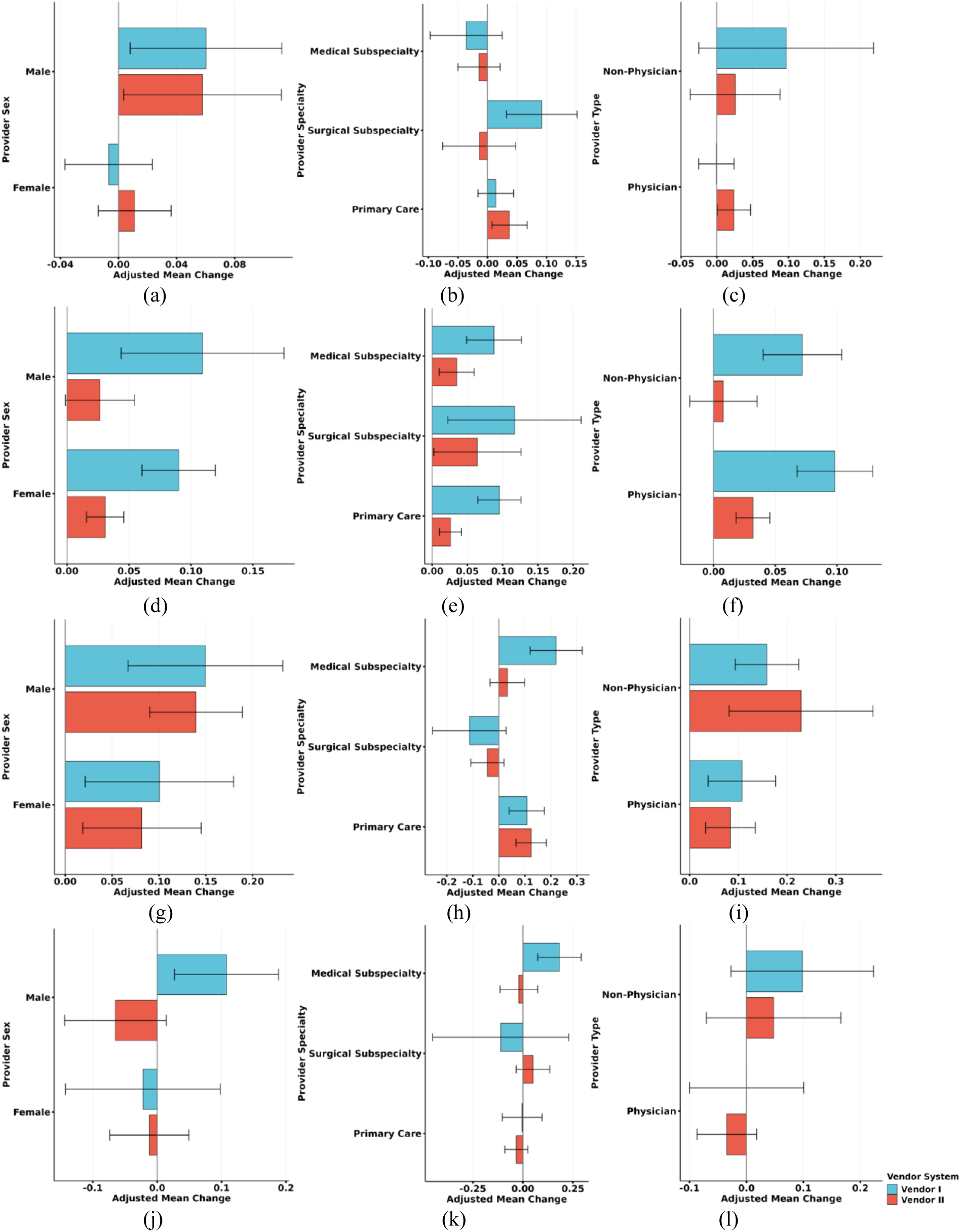
Medical term density for HPI section in (a) Sex (b) Provider Specialty (c) Provider type Medical term density for A&P section in (d) Sex (e) Provider Specialty (f) Provider type. MedReadMe for HPI section in (g) Sex (h) Provider Specialty (i) Provider type MedReadMe for A&P section in (j) Sex (k) Provider Specialty (l) Provider type.

For medical term density in the HPI section, Vendor I showed modest increases for males and surgical subspecialists (*P* = 0.024, 0.003), while other subgroups were nonsignificant. Vendor II had similar patterns, with small but significant increases for males, primary care, and physicians (*P* = 0.014–0.044). In the A&P section, Vendor I exhibited consistent and significant increases across almost all subgroups (*P* ≤0.016), whereas Vendor II showed smaller changes, with some subgroups (male and non-physician) not reaching significance.

For MedReadMe, which reflects readability of medical text, the HPI section showed significant improvements for most subgroups for both vendors, particularly provider sex, primary care, and provider type (*P* ≤0.003). Surgical subspecialists in Vendor I and Vendor II were exceptions with nonsignificant decrease. In the A&P section, significant MedReadMe increases were limited; Vendor I showed significant improvement only for males and medical subspecialists (*P* = 0.001–0.010), whereas Vendor II changes were mostly nonsignificant across subgroups.

Overall, the figure indicates that medical term usage and readability improvements were more pronounced in the HPI section, with variability across providers and vendors, while the A&P section showed smaller and less consistent effects.

Manual evaluation was performed to assess the consistency of human ratings across clinical notes. The overall inter-rater agreement demonstrated moderate reliability, with a two-way ICC of 0.62. Figure 8 compares manual human evaluations with automatic coherence and readability metrics across two vendors, Vendor I and Vendor II, under Ambient and Non-Ambient conditions, revealing consistent trends between human judgment and automated assessment. In the manual evaluation results shown in panels (a–c), Ambient conditions consistently achieve higher scores than Non-Ambient conditions for both coherence and readability. For the combined results in panel (a), Ambient speech attains HPI and A&P coherence scores of 3.29 and 3.42, respectively, compared to 2.97 and 3.19 in Non-Ambient settings, while readability similarly improves from 3.29–3.32 to 3.61–3.62. Vendor I-specific results in panel (b) show the strongest overall manual performance, with Ambient HPI readability reaching 3.80 and A&P readability 3.54, whereas Non-Ambient scores drop to 3.18 and 3.24. Vendor II results in panel (c) also favor Ambient conditions, particularly for A&P metrics, where coherence and readability reach 3.60 and 3.70, respectively.

**Figure 8.**
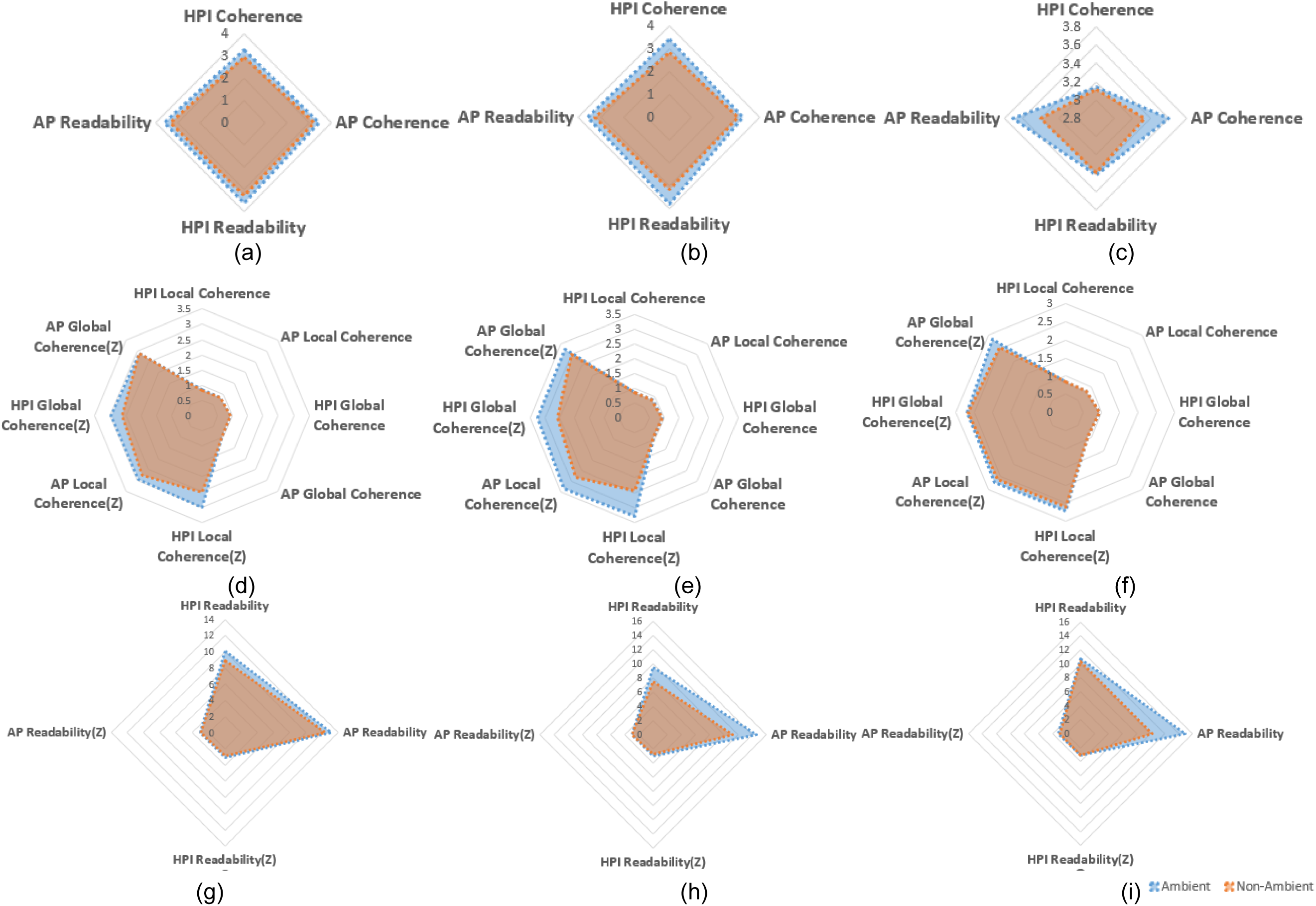
Manual and automatic evaluation of coherence and readability across vendors and sections for the subset. Manual ratings of HPI coherence, A&P coherence, HPI readability, and A&P readability for (a) both vendors, (b) Vendor I, and (c) Vendor II. Coherence metrics, including local and global coherence and their Z-score–normalized values (scaled to 1–5), for (d) both vendors, (e) Vendor I, and (f) Vendor II. Readability metrics and corresponding Z-score–normalized scores for (g) both vendors, (h) Vendor I, and (i) Vendor II

This trend is reinforced by the automatic coherence metrics shown in panels (d–f). For all vendors combined (panel d), Ambient conditions exhibit higher local and global coherence values for both HPI and A&P, with normalized Z-scores ranging from approximately 2.92 to 3.01, compared to 2.53 to 2.92 under Non-Ambient conditions. Vendor I (panel e) demonstrates the largest Ambient advantage, with normalized local coherence Z-scores increasing from 2.46–2.81 in Non-Ambient speech to 3.31–3.33 in Ambient speech, alongside corresponding gains in global coherence. Vendor II (panel f) shows more modest differences between conditions, though Ambient speech still consistently outperforms Non-Ambient speech across both local and global coherence measures.

Automatic readability results in panels (g–j) closely mirror the manual findings. For the combined dataset (panel g), Ambient readability scores are higher for both HPI (10.11 vs. 8.92) and A&P (13.13 vs. 12.33), with normalized Z-scores improving from approximately 2.90–2.98 to 3.04–3.07. Vendor I readability in panel (h) shows a clear Ambient advantage, particularly for A&P readability, which increases from 11.22 in Non-Ambient conditions to 14.62 in Ambient conditions. Vendor II readability in panel (i) similarly benefits from Ambient conditions, with A&P readability rising from 10.22 to 14.88 and normalized scores increasing from 2.92 to 3.16. Overall, the figure demonstrates strong agreement between manual and automatic evaluations and highlights the consistent negative impact of Non-Ambient conditions on both coherence and readability across vendors.

## 4. Discussion

This study provides a multi-dimensional examination of ambient clinical documentation, offering insights into how ambient intelligence reshapes the linguistic properties of clinical notes beyond basic lexical patterns. Rather than focusing on specific quantitative differences, our findings suggest that ambient documentation introduces systematic changes in how clinical information is expressed, organized, and standardized, reflecting both the affordances and constraints of current ambient AI systems.

Ambient documentation systems appear to standardize clinical language, reducing ambiguity and enhancing coherence. This effect likely arises from the models’ reliance on learned linguistic patterns and summarization templates, which favor explicit phrasing and syntactic consistency over idiosyncratic provider-specific expressions. Through translating conversational input into structured narratives, ambient systems may implicitly resolve underspecified references, normalize terminology, and reduce informal shorthand common in manually authored notes. At the same time, the observed increases in lexical complexity and readability indices indicate that ambient systems may prioritize completeness and formality over brevity or accessibility. While this enhances thoroughness and reduces misinterpretation, it may also increase cognitive load for downstream users, including trainees, interdisciplinary teams, and patients accessing open notes. Consistent with these quantitative findings, qualitative feedback revealed tension around information density: some clinicians felt that ambient AI omitted specific details, whereas others reported that it sometimes added too many details, making it harder to locate key information when reviewing prior notes. One primary care clinician noted that by “it could add too many details and make it harder to look back on previous notes and isolate the information that I need,” and another emphasized that excessive elaboration could make it difficult for future treating colleagues to quickly identify relevant content [39]. Importantly, the strong semantic alignment with non-ambient notes suggests that ambient documentation largely preserves clinical intent, functioning as a rephrasing and restructuring mechanism rather than altering clinical reasoning itself, a critical factor for trust and adoption.

### 4.1 Interpretation of Specific Linguistic Metrics

To mitigate clinician documentation burden, healthcare systems have long relied on dictation technologies and medical scribes, each of which changes how clinical language is produced rather than how it is fundamentally structured [40]. Prior studies show that scribed notes tend to be longer largely because of templated and copied content, contributing to note bloat without necessarily increasing informational value [41]. Similarly, work comparing voice-dictated and typed notes demonstrates that documentation modality shapes linguistic characteristics beyond length, including vocabulary choice, sentence structure, lexical diversity, part-of-speech distributions, and error profiles [42]. These findings suggest that how documentation is generated influences not only efficiency but also the linguistic form and interpretability of clinical notes [43]. Ambient AI builds on these modalities but differs in a critical way: rather than merely transcribing speech or offloading typing, it actively reconstructs clinical narratives through summarization, normalization, and discourse planning.

This distinction helps explain observed changes in sentence- and clause-level structure. Ambient systems ingest conversational input and reorganize it into clinically coherent prose, which can lead to greater syntactic embedding and denser information packing. The sentence structure in ambient notes is more complicated, having more pronounced effects in the HPI section, where conversational speech is transformed into integrated narrative prose. For example, in one case of Vendor I, descriptions of carpal tunnel syndrome include symptom characterization, activity triggers, and treatment instructions all within unified sentences (“Presents with tingling and sensitivity in the hand, particularly after heavy lifting and physical work… Discussed the use of a wrist splint… Emphasized the importance of continuous use for two weeks, then nightly use”). Longer sentences or clauses in this context do not necessarily indicate reduced clarity; instead, they often reflect consolidation of fragmented, telegraphic utterances common in manually typed or dictated notes into unified, context-rich statements. Dependency distance patterns further support this interpretation: increased syntactic dispersion in ambient-generated HPI sections likely reflects deliberate narrative reconstruction that links symptoms, timelines, and modifiers into integrated structures, rather than inefficiency or redundancy. In contrast, shorter dependency spans are more consistent with templated phrasing, which prioritizes brevity and standardization over narrative richness.

Part-of-speech and entropy patterns suggest that ambient systems perform adaptive, section-sensitive linguistic normalization rather than uniform stylistic flattening. In narrative contexts such as the HPI, increased syntactic diversity and greater use of functional categories (e.g., determiners, conjunctions, verbs) indicate expansion and scaffolding of patient stories into more grammatically explicit, cohesive prose, reducing telegraphic phrasing and enhancing connective structure. In contrast, in the A&P, particularly for Vendor I, the shift toward noun-dominant, lower-entropy text reflects structural consolidation and content densification, emphasizing diagnostic reasoning and management planning in a more compressed, standardized format. Vendor II shows a milder version of this effect, maintaining variability rather than strongly compressing structure. Different vendors are affected by different training data, template design, and fine-tuning strategies, which may explain why POS entropy decreases in Vendor I’s A&P but not consistently in Vendor II. Overall, ambient systems appear to redistribute linguistic variability according to section function, simultaneously elaborating narrative documentation and stabilizing evaluative content, thereby reducing idiosyncratic provider variation while promoting institutional consistency and downstream usability.

Readability findings are best understood through the concept of medical sublanguage rather than general-audience comprehension. Ambient systems more frequently substitute precise clinical terminology for lay paraphrases, reflecting optimization for professional communication rather than patient-facing clarity. For instance, Vendor I uses terms such as “irbesartan” and “Metformin XR” with specific dosing instructions, while Vendor II similarly specifies “atorvastatin 20 mg daily” and detailed laboratory follow-up plans.

Because MedReadMe is calibrated using annotations from non-medical readers, specialized terms are penalized as increased processing effort, even when clinicians perceive them as clearer, more accurate, and less ambiguous. This explains why ambient documentation appears more complex under MedReadMe despite being judged by healthcare providers as more readable and more suitable for billing, legal, and medico-legal purposes. In other words, ambient AI shifts documentation toward a professional sublanguage optimized for clinical precision rather than general accessibility.

The section-specific nature of these effects is also consistent with workflow realities. The HPI is inherently narrative and conversational, making it particularly amenable to AI-driven restructuring, temporal alignment, and reference resolution. Ambient systems can therefore add conceptual density and coherence by transforming speech into organized clinical storytelling. In contrast, the A&P section is already highly structured, relies heavily on standardized templates, and is less dependent on conversational input, thus ambient systems reinforce and standardize this structure rather than fundamentally expand it. Both vendors employ structured A&P templates that prioritize diagnosis-first organization, followed by diagnostic reasoning and treatment plans. Ambient documentation typically captures the diagnosis first, then organizes relevant supporting information under each condition, and finally specifies treatment or follow-up plans. For example, Vendor I frequently generates blocks such as “Primary diagnosis – Treatment plan – Follow-up,” while Vendor II presents structured “Orders” lists embedded within each diagnosis. This contrasts with non-ambient notes, where different providers often use idiosyncratic ordering, mixed narrative and bullet formats, or abbreviated problem lists. Ambient notes therefore display a more uniform structural style across encounters; providers also tend to engage ambient tools less when authoring A&P content. As a result, ambient AI has limited opportunity and limited necessity to alter linguistic form in this section.

Improvements in linguistic predictability and coherence further reflect this normalization process. Through mapping conversational language onto clinically grounded patterns, ambient systems reduce variability and ambiguity in word choice, aligning notes more closely with domain-specific language models such as BioGPT. At the same time, gains in coherence are strongest where discourse structure matters most within narrative sections, while remaining attenuated in list-based or problem-oriented content. Finally, heterogeneity across providers and encounters likely reflects variation in speech patterns, documentation habits, and clinical complexity, which interact with how ambient AI summarizes and structures input rather than being overridden by it.

The heterogeneous effects observed across provider and clinical subgroups likely reflect differences in documentation style, workflow integration, and clinical narrative demands rather than differential performance of the ambient systems themselves. Provider specialty appears to be a major driver: narrative-intensive fields such as primary care, pediatrics, psychiatry, and hematology rely heavily on longitudinal histories, symptom evolution, and contextual detail, creating greater opportunity for ambient AI to restructure conversational input into more coherent, information-dense prose. In contrast, procedure-oriented or highly protocolized specialties (e.g., surgery, urology, oncology) often document using standardized templates and concise problem lists, limiting the extent to which ambient systems can meaningfully alter syntactic structure, coherence, or readability. Differences by provider type likely reveal baseline documentation practices and role-specific note ownership: advanced practice providers and physicians who author comprehensive encounter notes may benefit more from ambient narrative reconstruction, whereas certain allied health or support roles tend to produce shorter, task-focused documentation that is less amenable to linguistic transformation. The minimal and inconsistent effects observed by provider sex suggest that documentation changes are driven more by professional role and clinical context than by demographic factors. Together, these subgroup patterns indicate that ambient documentation amplifies existing documentation norms, enhancing structure, coherence, and precision where narrative complexity is inherent, while exerting limited influence in already standardized or constrained documentation settings.

**Table 6.** Summary of findings.

| Domain | HPI | A&P | Vendors |
| --- | --- | --- | --- |
| Syntactic complexity | • Increased sentence length<br>• Increased clause length<br>• Increased dependency distance<br>• Increased POS entropy | • Decreased clause length<br>• Decreased dependency distance | • Effects stronger and more consistent for Vendor I<br>• Vendor I showed clear A&P simplification<br>• Vendor II showed minimal A&P sentence length change<br>• Vendor I: POS entropy increased in HPI but decreased in A&P<br>• Vendor II: POS entropy increased in both sections<br>• Vendor I showed stronger clause-level increases across sex, specialty, and physician groups |
| Lexical ambiguity | • Significant reduction<br>• Reductions consistent across sex and primary care<br>• Stronger effects in HPI than A&P | • Significant but smaller reduction | • Vendor I showed more consistent reductions across subgroups<br>• Vendor II effects less consistent in surgical and some subspecialties<br>• Non-physician reduction stronger in Vendor II |
| Linguistic variability | • Overall reduction<br>• Reduced variability and more standardized language | • Substantial reduction<br>• Stronger and more consistent than HPI | • Vendor I showed larger and more consistent reductions<br>• Vendor II HPI reductions inconsistent with BioGPT<br>• GPT-2 reductions more stable than BioGPT<br>• Vendor I effects consistent across sex and primary care |
| Discourse coherence | • Increased local coherence<br>• Increased global coherence<br>• Increased shuffled local coherence<br>• Increased shuffled global coherence | • Small changes in global coherence | • Vendor I showed consistent improvements across nearly all subgroups<br>• Vendor II A&P global coherence slightly decreased<br>• Vendor II A&P shuffled measures often nonsignificant<br>• Vendor I improvements stronger in primary care and physicians<br>• Shuffled difference close to zero for most groups |
| Readability | • Increased word count<br>• Increased complex word count<br>• Increased sentence count<br>• Improved MedReadMe readability | • Increased complex word count<br>• Increased medical term density | • Word count increases larger for Vendor I in HPI<br>• A&P word count significant mainly for Vendor II<br>• Medical term density increases stronger in Vendor I A&P<br>• MedReadMe improvement consistent in HPI but not A&P<br>• HPI length effects stronger across sex and provider type<br>• A&P structural changes more variable across vendors |

### 4.2 Novel Contributions and Limitations

This study introduces several novel contributions to the evaluation of ambient AI–assisted clinical documentation. First, it adopts a holistic linguistic evaluation, simultaneously examining ambiguity, coherence, complexity, and readability, providing a comprehensive view of the changes introduced by ambient AI. In addition, a subset of 50 paired clinical notes was manually evaluated for readability and coherence to contextualize and validate the automated metrics. Second, within-patient comparisons allow us to minimize confounding by patient-level factors, isolating the effects attributable to the documentation system itself. Third, by exploring variation across note sections, vendors, and provider characteristics, we capture the real-world heterogeneity inherent in clinical documentation. Finally, by connecting linguistic properties to their potential downstream impact on information retrieval, decision support, and secondary data use, the study bridges AI-driven documentation changes with meaningful clinical applications.

Several limitations should be acknowledged. First, this study is based on data from a single healthcare organization, which may limit the generalizability of findings to other institutions. Second, while we assessed multiple linguistic and semantic properties, we did not directly evaluate clinical accuracy, patient outcomes, or workflow efficiency, which remain important areas for future investigation. Third, our analyses do not capture provider perceptions, editing behaviors, or trust, factors that influence adoption and real-world use of ambient documentation systems. In addition, the distribution of provider roles was imbalanced, with relatively few nursing-authored notes compared to notes authored by attending physicians and advanced practice providers, which may have disproportionately influenced the observed documentation-related gains and limits the interpretability of role-specific effects. Finally, ambient intelligence platforms continue to evolve, so observed patterns may reflect characteristics of the current generation of models rather than enduring features of such systems.

### 4.3 Implications for Clinical Integration

The findings of this study have several practical implications for the adoption and integration of ambient documentation technology in clinical settings while highlighting the challenges that must be addressed to achieve meaningful use. Ambient systems improve linguistic predictability, coherence, and readability, particularly in narrative-driven sections such as HPI, while preserving clinical intent. Through reorganizing conversational input, normalizing terminology, and resolving underspecified references, these systems help providers produce clearer, more standardized notes without altering diagnostic reasoning. This can reduce misinterpretation, enhance documentation quality, and facilitate downstream applications such as clinical decision support, quality measurement, and research. Moreover, improvements in readability and syntactic consistency may enhance accessibility for interdisciplinary teams, trainees, and patients accessing open notes, while denser sentence structures in some sections highlight the need to balance thoroughness with cognitive load. Thoughtful interface design, user training, and adaptive presentation formats can support comprehension without overwhelming end users, ensuring the benefits of ambient systems are realized in practice.

The heterogeneous effects observed across provider characteristics, specialties, and visit types underscore the importance of context-aware implementation strategies. Organizations should consider tailoring system configuration, providing usage guidance, or calibrating models to accommodate differences in dictation style, note complexity, and clinical context. Such targeted implementation can help ensure consistent improvements in readability, coherence, and syntactic structure across diverse clinical scenarios. At the same time, while A&P and other structured sections show limited gains, maintaining content fidelity remains critical. Ambient documentation should complement rather than replace provider oversight, with integrated real-time editing workflows, review mechanisms, and feedback loops to prevent omission or misrepresentation of critical information, support cognitive efficiency, and maintain trust in documentation quality.

Finally, these findings highlight the need for ongoing monitoring and evaluation of ambient systems post-deployment. As AI platforms continue to evolve, linguistic outputs, section-specific performance, and subgroup-level effects may shift, requiring continuous assessment to ensure equitable and clinically meaningful benefits across all provider groups and patient populations. Through combining careful system design, context-sensitive implementation, and iterative evaluation, healthcare organizations can leverage the strengths of ambient AI while addressing potential risks, supporting provider reasoning, and improving the overall quality and usability of clinical documentation.

## 5. Conclusions

This study provides a systematic and comprehensive study of how ambient intelligence reshapes clinical documentation at the linguistic level while maintaining clinical semantics. It highlights the potential for Ambient intelligence to enhance clarity, consistency, and quality in healthcare communication and coordination, providing a foundation for its meaningful use and integration into clinical practice.

## Acknowledgments

This article was partially supported by NIH/NLM under Award Numbers 1R01LM014239 & T15LM007092 and NIH/NIA under Award Numbers R01AG080429.

## Ethics Approval and Consent to Participate

Not applicable.

## Author Contribution

**Yiming Li**: Conceptualization; Methodology, Formal analysis, Software, Investigation, Data Curation, Visualization, Writing - Original Draft **Huiyuan Zhou:** Formal analysis, Visualization, Writing - Review & Editing **Suzanne Blackley:** Data Curation **Joseph M. Plasek:** Writing - Review & Editing **Zhuoyang Lyu:** Visualization **Wei Zhang:** Data Curation **Jacqueline You:** Resources, Funding acquisition, Writing - Review & Editing **Amanda Centi:** Resources **Rebecca Mishuris:** Resources **Jie Yang:** Resources, Writing - Review & Editing **Li Zhou:** Writing - Review & Editing, Supervision, Project administration, Funding acquisition

## Data Availability

The data generated and analyzed during the current study are not publicly available due to organizational patient privacy policies. Any request to access the Data will need to be reviewed and approved by Mass General Brigham. Researchers will need to provide evidence of IRB approval for their study. For the approved studies, data will be released via a Data Use Agreement.

